# Modelling the influence of the duration of effectiveness and time taken to implement vector control interventions for vector-borne diseases

**DOI:** 10.64898/2026.09.30.26364329

**Authors:** Nilani Chandradeva, Andrew C. Glover, Charles Whittaker, Tom Brewer, Joseph D. Challenger, Hannah C. Slater, Ellie Sherrard-Smith, Thomas S. Churcher

## Abstract

Implementing vector control interventions against infectious diseases is operationally challenging, with delivery and population uptake taking considerable time within a community. Studies have suggested that the slow mass drug administration (MDA) of the endectocide, ivermectin, may have contributed to the lower epidemiological impact against malaria observed in recent cluster-randomised controlled trials, although evidence supporting this hypothesis is lacking. It is unclear whether interventions which have short but highly effective durations of entomological impact are more impactful than tools which kill the same number of mosquitoes but over a longer period. Transmission dynamics mathematical models can be used to explore the trade-offs between product profiles and investigate how different implementation periods could influence disease control. First, we use a simple vector-borne disease compartmental model to explore the epidemiological impact of killing the same number of mosquitoes over 10, 30 or 90 days. We find that for different mosquito-killing periods, the total number of cases averted is similar, although the observable impact may depend on when efficacy is assessed. Secondly, we adapted an existing malaria transmission model and simulated the fast and slow deployment of an effective endectocide MDA campaign in different seasonal settings. In perennial and seasonal settings and for a given coverage, slow MDA deployment averts more clinical cases of malaria than a fast synchronised distribution (completed in a single day, although the difference was modest). The benefits of the intervention depend on how well-timed a deployment is relative to the peak of the transmission season, with the efficacy of an ivermectin-like endectocide MDA being up to approximately 42% lower if started late. The work indicates that the time taken to complete MDA campaigns is unlikely to drive major differences in the effectiveness of short-lived endectocides and showcases how transmission dynamics models can be used to support epidemiological trial interpretation.

**Author summary:** Mathematical models can be used to help guide the development and implementation of control interventions against infectious diseases and explore how real-world complexities can impact disease control. Previous modelling work indicated that the mass drug administration (MDA) of the mosquito-killing drug, ivermectin, could substantially reduce malaria transmission, assuming eligible individuals due to receive the drug do so on the same day. Logistical constraints mean that MDA campaigns are often slower to implement, although the epidemiological impact of this is unclear. We develop a mathematical modelling framework to explore how the speed at which an intervention is deployed, or its period of effectiveness, influences vector-borne disease dynamics. Results suggest that spreading the impact of an intervention over a longer period provides slightly greater protection. Slower MDA deployment of an ivermectin-like drug is generally more efficacious, although impact is diminished in areas with seasonal mosquito abundance if it is implemented too late in the transmission season.

## Introduction

Killing mosquitoes is one of the most effective ways of reducing the burden of diseases that they transmit. The most substantial gains in reducing malaria transmission in Africa can be attributed to the wide scale deployment of insecticide treated nets (ITNs) and indoor residual spraying (IRS) [1]. The number of malaria cases remains intolerably high, so new vector control tools are urgently needed, particularly those which target residual transmission, defined as transmission still occurring when indoor interventions (ITNs and IRS) are working optimally [2, 3]. Gaps in protection arise from several factors including complex mosquito biting behaviours which reduce insecticidal exposure (crepuscular biting behaviours [4, 5], diversion to feed on alternative hosts, such as livestock, and potential avoidance of insecticide-treated spaces [6]), insecticide resistance that reduces the epidemiological impact of ITNs [7], or the inability to achieve and maintain high use of control tools [8].

Up to now, vector control tools have been relatively long lasting, with ITNs designed to remain effective for three years and IRS intended to maintain effectiveness throughout the transmission season. New tools under development have a much shorter period of effectiveness. For example, spatial emanators which emit volatile insecticide have recently been recommended by the World Health Organisation for indoor use alongside ITNs [9], with individual devices remaining effective from as little as a month, though other devices can be effective for up to a year [10]. The mass-fogging applications of insecticides has been extensively used in higher income areas, though its killing ability may only last for a few days depending on method of application and climate variables, and there is conflicting evidence on the impact of the intervention on non-target insects [11, 12]. Endectocides are a promising type of drug; when administered to humans or livestock, they enter the bloodstream and mosquitoes are killed by the drug if they feed on the individual whilst the blood-drug concentrations are at a lethal dose. The period of action of endectocides varies between products and dosing regimens. Ivermectin is the most widely studied candidate endectocide for malaria control, and thought to have a killing effect in *Anopheles* mosquitoes for approximately 23 days at a dose of 300 *µ*g per kilogram of bodyweight, administered on 3 consecutive days (3×300 *µ*g/kg), although the magnitude of the effect depends on the blood-drug concentration, which wanes over the course of this 23-day period [13]. However, the epidemiological impact of trade-offs between entomological effectiveness, for different products, and period of activity, is unknown. For example, it is unclear whether a product which has a higher entomological efficacy but a short period of activity (e.g. kills a target number of mosquitoes over a short period of time) would have a larger epidemiological impact than one which has less of an entomological impact but over a longer period of activity (e.g. kills a target number of mosquitoes over a relatively longer period of time).

Mathematical modelling can be used to predict the impact of novel vector control products in different settings. For novel control tools, models can be particularly useful to explore the theoretical epidemiological impact of interventions with different target product profiles and modes of action that can support the developmental process [14–17]. Similarly, they could be used to investigate how different real-world deployment scenarios could influence the efficacy of a novel tool. Previous modelling work has predicted that the mass distribution of ivermectin could substantially reduce malaria burden in different malaria endemicity settings [15]. This model assumed that, in a cluster-randomised controlled trial (cRCT) setting, all individuals receiving the mass drug administration (MDA) in a cluster were treated simultaneously. However, such synchronised MDA of ivermectin in recently completed cRCTs was not feasible, with MDA campaigns taking between approximately 9 and 20 days to complete [18, 19]. For example, the MATAMAL trial (ClinicalTrials.gov registration: NCT04844905) in the Bijagos archipelago, Guinea-Bissau, reported that it took, on average, 9.4 days (SD = 2.3), to distribute 80% of the target coverage of MDA to the intervention group and 9.1 days (SD = 2.0) for the corresponding placebo coverage in the control group, and it has been suggested that the prolonged nature of the MDA campaign could explain the null trial result of this trial [18]. The BOHEMIA trial (ClinicalTrials.gov registration: NCT04966702) was conducted in two settings (in Mozambique and Kenya) [20]. In Mozambique, the implementation of the campaign was influenced by cyclone Gombe, resulting in a later start of the campaign and it taking longer to complete the campaign (on average, campaigns took 18 days to complete) [19]. This trial had a null result. On the other hand, the MDA campaigns in Kenya took on average 10 days to complete and this trial reported a significant difference in malaria infection incidence in children aged 5-15 years between trial arms [21], leading to the hypothesis that faster MDA campaigns are more impactful than slower campaigns [19]. However, there were many other differences between these trial sites, making it difficult to attribute the time taken to implement the MDA as the driver of these results. In real-world settings, when distributions would be conducted by national programmes rather than trial teams, this may take even longer. Funding MDA campaigns to be synchronised such that they can be completed in short time periods would be extremely expensive for national programmes, potentially limiting the affordability of this intervention at a time of major uncertainty in global health budgets. Therefore, understanding the epidemiological impact of the time taken to achieve population coverages and kill mosquitoes will be important for the operational planning of MDA campaigns for other endectocides under development.

It is infeasible to empirically test how different product profiles or deployment scenarios influence the epidemiological impact of different vector control tools such as endectocides. Instead, here we use transmission dynamics models to explore how interventions with different periods of effectiveness (analogous to varying the time taken to distribute short-acting vector control products) influences its epidemiological impact. First, we use a simple generic vector-borne disease transmission model to simulate the killing of a target number of mosquitoes over different time periods, and compare transmission dynamics and intervention efficacy in these scenarios. We then explore the specific situation of how the time taken to complete MDA campaigns of an ivermectin-like endectocide may impact malaria control.

## Materials and methods

### Simulating slow and fast killing of mosquitoes using a simple SEIRS-SEI model

A deterministic compartmental model of a generic vector-borne disease (for example, dengue virus or malaria) was developed to explore the influence of killing a target number of mosquitoes within different time periods (10 days, 30 days or 90 days). The human population is assumed to be either Susceptible (*S*_*h*_), Exposed (*E*_*h*_, exposed but not infectious), Infectious (*I*_*h*_) or Recovered (*R*_*h*_) whilst the vector population is either Susceptible (*S*_*v*_), Exposed (*E*_*v*_) or Infectious (*I*_*v*_). The compartmental model is illustrated in Fig 1 with the equations written in the Supporting Information. It is parameterised for a malaria-like disease using values outlined in S1 Table, and was implemented in the *odin* package (R version 4.4.1) [22, 23].

**Fig 1.**
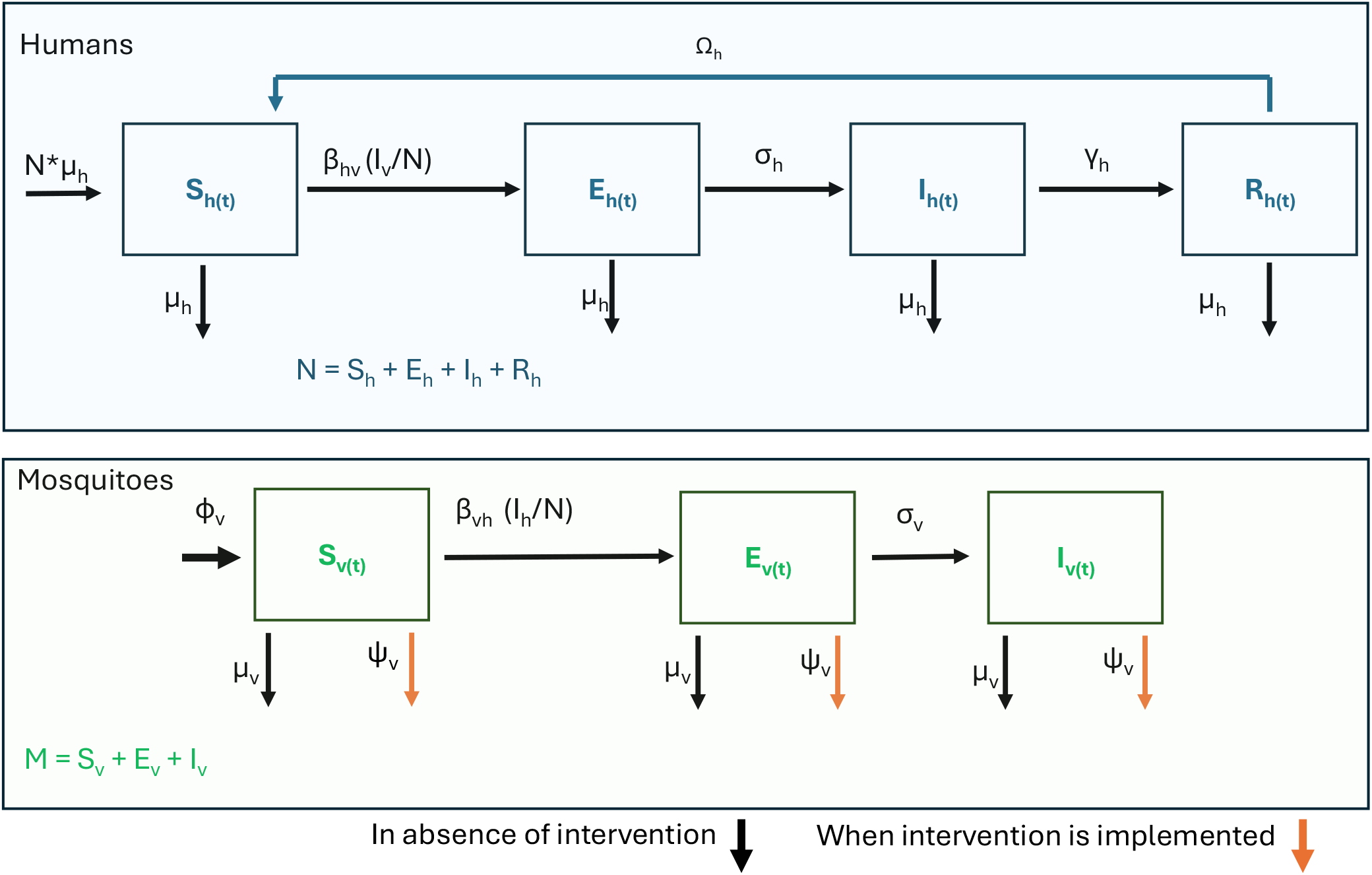
Simple SEIRS-SEI model for malaria transmission. Infection in humans is modelled using an SEIRS (*S*_*h*_: susceptible; *E*_*h*_: exposed; *I*_*h*_: infectious; *R*_*h*_: recovered) framework. Human immunity wanes at rate Ω_*h*_. *β*_*hv*_ is the effective daily vector to human transmission rate; *σ*_*h*_ is the intrinsic incubation period and *γ*_*h*_ is the human infectious period. Infection in mosquitoes is modelled using an SEI framework in mosquitoes (*S*_*v*_: susceptible; *E*_*v*_: exposed; *I*_*v*_: infectious). When the intervention is implemented, mosquitoes experience a mortality rate, *ψ*_*v*_, in addition to the background (natural) mosquito mortality rate, *µ*_*v*_. Mosquitoes emerge at rate *ϕ*_*vi*_, where i = 1 refers to a constant emergence rate (Equation 1) and i = 2 refers to an emergence rate governed by a logistic growth model (Equation 2). *β*_*vh*_ is the effective daily human to vector transmission rate and *σ*_*v*_ is the extrinsic incubation period. M is total number of mosquitoes and N is the total number of humans. All model parameter values are given in S1 Table.

The model was parameterised so that 1,900 mosquitoes, in a population of 2,000 mosquitoes at endemic equilibrium, were killed in each of the different time periods. The additional mortality rate required to kill the target number of mosquitoes in each time period is denoted as *ψ*_*v*_. The method used to estimate *ψ*_*v*_ for each time period is described in the Supporting Information and varied depending on the assumption on the emergence rate of adult mosquitoes. The model parameterisation is shown in S1 Table which includes a vector-host ratio of 2 mosquitoes per person. A sensitivity analysis was conducted in which 190,000 mosquitoes, in a population with 200,000 at endemic equilibrium, were killed over the different time periods (increasing the vector-host ratio to 200 mosquitoes per person).

Mosquito populations are regulated by different mechanisms, including negative density-dependent processes, which reduce population growth at higher densities and ensure a stable endemic equilibrium. The type of density dependence influences how mosquito populations rebound following perturbation and could have epidemiological implications for the difference between short and longer acting vector control interventions. Here the population is assumed to be regulated by within-species competition of insect larvae for resources such as food, which causes increased mortality at high larval densities, and by environmental carrying capacities which limit the total number of vectors that an environment can sustain (for example, the availability and size of larval breeding sites) [24]. We explored the influence of different assumptions on the adult emergence rate (*ϕ*_*vi*_) on the epidemiological impact of the interventions with different mosquito-killing durations. One assumption is that the emergence rate is constant (*ϕ*_*v,i*=1_) and therefore the number of new adult mosquitoes is independent of the number of mosquitoes killed:

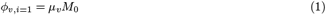

where *M*_0_ denotes the number of mosquitoes at endemic equilibrium (initial population size) and *µ*_*v*_ is the natural mosquito mortality rate. We also explore the influence that a logistic growth model has on adult emergence (*ϕ*_*v,i*=2_), through affecting the number of mosquitoes that the environment can support:

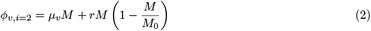

where *M* is the adult female mosquito population at a given time point and *r* is the intrinsic growth rate (here, it is assumed that *r* = *µ*_*v*_).

The entomological efficacy of the intervention is measured as the percentage of mosquitoes killed (*D*, equations shown in the Supporting Information) during the time since the intervention started (*s*), compared to the total number of mosquitoes that have emerged within that period. The entomological efficacy for each mosquito-killing period is measured at 10 days, 30 days and 90 days and explored under both assumptions of adult mosquito emergence (*ϕ*_*v,i*_) as follows:

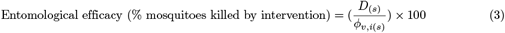

In trials, the efficacy of interventions can also be estimated using cross-sectional prevalence surveys and measuring the percentage reduction in prevalence between the control and intervention arm. Therefore, we additionally estimate efficacy in prevalence (for each emergence rate scenario) as follows:

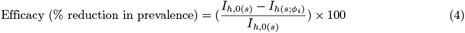

The reduction in cases, due to the intervention, was estimated by comparing the cumulative number of cases over each of these time periods (*C*_*h*_) to a counterfactual scenario of no interventions (*C*_0_) at these different time points (10 days, 30 days or 90 days since the start of the intervention (*s*)) to calculate the cases averted (Δ*C*) over each time-frame, *s*, under each emergence rate assumption:

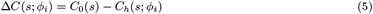

The relative efficacy of each MDA deployment scenario was then additionally assessed from the percentage

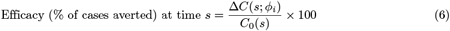

A sensitivity analysis was conducted to determine the influence of endemicity (determined by increasing the vector-host ratio to 200) on the efficacy of the different interventions.

### Simulating endectocide mass drug administration (MDA) with synchronised and slow deployment

We used a deterministic version of a validated mechanistic *Plasmodium falciparum* malaria transmission model [25, 26] to further explore the influence of the time taken to implement MDA campaigns of an ivermectin-like endectocide on malaria transmission dynamics and epidemiological impact. We explored these dynamics in a high transmission intensity setting (in the absence of interventions, an individual receives on average 100 infectious bites per year). Seasonality in transmission was captured using a Fourier series which adjusts mosquito emergence in response to satellite-derived rainfall estimates. The model structure is described in detail in the Supporting Information. In this model, density dependence was driven by the vector-host ratio in the absence of interventions (but affected by seasonality) and the mortality rate (in the absence of interventions), i.e. the number of adult mosquitoes emerging into the adult population is independent of the number of mosquitoes killed by interventions.

Using the mass drug administration (MDA) of an ivermectin-like endectocide as an example, we compared the epidemiological impact between synchronised campaigns, in which all individuals due to receive the MDA (the target coverage amongst the eligible population) do so at the same time (an idealistic approach, previously modelled [15, 27, 28]), to slower campaigns in which it takes 10 days or 20 days to distribute the MDA campaign to all individuals due to receive it. The structure of the human malaria model is shown in S3 Fig and all equations are shown the Supporting Information. In this model, the killing effects of ivermectin were captured using hazard ratios (HR), which quantify how much more likely a mosquito is to die if it feeds on an ivermectin-treated person compared to someone who is untreated (If the HR = 1, mosquitoes are equally likely to die if they have fed on treated or untreated individuals). Following a phase II clinical trial (the IVERMAL trial) [13], a 3×300 *µ*g/kg dosage of ivermectin is assumed to result in HRs that are greater than 1 for 23 days, and the highest HR is achieved on the third day since the first dose [27]. The HR curve, with HR values for each day since the first dose, is shown in S4 Fig.

We simulated the slower MDA deployment scenarios such that a target coverage (the coverage of the intervention amongst the population that are eligible (*p*_E_) to receive an ivermectin-like endectocide, *c*_target_) of 70% is achieved over three equally inter-spaced distributions. This is similar to treating different groups within the target population over time, which may be essential in a trial setting if the target population or cluster is too large to treat everyone on the same day. As the day on which these groups start the MDA is different, the HR in each group will differ on a given day. Therefore, the proportion of the covered groups with a blood concentration of endectocide that is lethal to mosquitoes (*K*_HR*>*1_, i.e. with a hazard ratio greater than 1) changes over time. Thus, the effective coverage of the ivermectin-like endectocide in the population also changes over time. To capture this, we introduce a time-varying parameter for the proportion of the population with a lethal dose of the endectocide (*Q*_lethal_). It is assumed that all individuals eligible to receive the MDA are over 5-years-old (*a*_min_), following safety criteria for ivermectin in recently completed cRCTs [20, 29, 30], for a population with a median age of 21-years-old (*a*_avg_), as assumed by the malaria transmission model [25]. We assumed that the maximum age (*a*_max_) is 90-years-old.

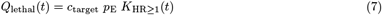

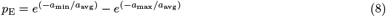

To account for changes in magnitude of the population-level hazard ratios over time (t) caused by the slower MDA deployment scenarios, we take, on a given day, the mean hazard ratios across each of the inter-spaced distributions that are greater than 1 (Equation 9, where *i* refers to the number of distribution groups and in this instance, *i* = 3). This assumes that in each mosquito feeding attempt, mosquitoes do not have a preference for a particular treatment group, or untreated individuals, and that if a mosquito feeds on individuals in multiple treatment groups, the hazard ratio it experiences corresponds to that of the first bloodmeal containing the endectocide. These hazard ratios are passed into the adapted model and used to elevate the mortality rate of mosquitoes.

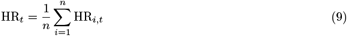

We simulated monthly synchronised and slow deployment scenarios for an ivermectin-like endectocide, for three months. Slow deployment scenarios took either 10 days or 20 days to complete. For each scenario, the corresponding values of *Q*_lethal_(*t*) and HR_*t*_ can be found in S5 Fig. These simulations were conducted for both a perennial and highly seasonal setting, using Fatick, Senegal, as an example for the latter. We modelled an additional scenario whereby the MDA started 60 days later than planned, at the time of peak clinical incidence in children under 5-years-old in the seasonal setting. Such disruptions to the timing of starting an MDA in relation to peak rainfall can occur due to logistical delays or if, for example, the rains start earlier than usual.

A sensitivity analysis was conducted to determine the influence of transmission intensity (by exploring the influence of the MDA deployment scenarios in a low transmission setting in which there are, on average, 2 infectious bites per person per year), and the coverage of the endectocide (between 50% and 90% coverage amongst the eligible population) on the epidemiological impact of MDA. Epidemiological impact is measured in three ways: cumulative clinical cases (*C*) averted in under 5-year-olds for 6 months from the point of the first MDA (as for the BOHEMIA trial [20]), all-age slide prevalence of *P. falciparum* (*Pf* PR) at 4 weeks after the last MDA (as for the MATAMAL trial, although in that trial, qPCR prevalence is used instead of slide prevalence [29]) and clinical cases averted in children under 5-years-old (*C*) for 1 year from the start of the first MDA, and comparing these (as a relative difference %) to a counterfactual scenario of no interventions (for cases averted, Equation 10; for reduction in *Pf* PR, Equation 11).

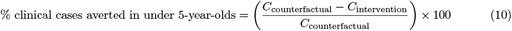

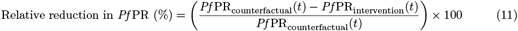

The analysis was conducted using R version 4.4.1. All scripts required to recreate the results of the analysis are available at: https://github.com/nilani-chandradeva1/control-intervention-speed.

## Results

### Epidemiological impact of killing the same number of mosquitoes over short or long periods

A simple SEIRS-SEI transmission model of a vector-borne disease was parameterised here to broadly simulate malaria transmission in an endemic, perennial setting. (The model structure and parameterisation are described in detail in the Methods section and Supporting Information.) The model was developed to explore the entomological and epidemiological impact of killing the same number of adult female mosquitoes over different time periods, assuming the emergence of adult mosquitoes is either governed by a logistic growth model or is constant over time. In this analysis, 1,900 mosquitoes (in a setting with an initial population size of 2,000 adult mosquitoes) were killed over 10 days, 30 days or 90 days (Fig 2A). The differences in the total number of mosquitoes over the different time periods of interest are shown in Table 1 (Equation 3). Regardless of the assumption on adult mosquito emergence, the highest percentage of mosquitoes killed by the different mosquito-killing implementation periods is for the 10-day killing period, when efficacy is measured 10 days since the start. However, by the time that all dynamics have returned to equilibrium (approximately 250 days since the start of the intervention, Fig 2A-E), a similar percentage of mosquitoes have been killed, regardless of the mosquito-killing period. These findings hold at a higher vector-host ratio (S2 Table). Fig 2B shows that the rate of rebound in mosquito population dynamics varies according to the assumption on adult mosquito emergence; if constant adult emergence is assumed, then the adult mosquito population returns to an endemic state faster than if a logistic growth model is assumed.

**Table 1.** Summary of the number (and percentage) of mosquitoes killed by the interventions with different mosquito-killing periods (10 days, 30 days or 90 days), as predicted by a generic SEIRS-SEI model of vector-borne disease transmission in which there are approximately 2 mosquitoes per person. The percentage of mosquitoes killed by the intervention was calculated by tracking the total number of mosquitoes killed by the intervention (the D compartment) compared to total number of mosquitoes to have emerged into the population (in the counterfactual no-intervention scenario) from the start of each mosquito-killing period to 10 days, 30 days, 90 days and 250 days later (All dynamics have returned to equilibrium by 250 days after the start of the mosquito-killing started). By each of these time points, 2,000, 6,000, 18,000 and 50,000 mosquitoes have emerged into the population, respectively.

| Mosquito-killing period (days) | Adult mosquito emergence assumption | N (%) of mosquitoes killed by intervention since it started |  |  |  |
| --- | --- | --- | --- | --- | --- |
|  |  | 10 days | 30 days | 90 days | 250 days |
| 10 | Constant rate | 1,900 (95.0) | 1,900 (31.7) | 1,900 (10.6) | 1,900 (3.8) |
|  | Logistic growth model | 1,900 (95.0) | 1,900 (31.7) | 1,900 (10.6) | 1,900 (3.8) |
| 30 | Constant rate | 705 (35.2) | 1,900 (31.7) | 1,900 (10.6) | 1,900 (3.8) |
|  | Logistic growth model | 742 (37.1) | 1,900 (31.7) | 1,900 (10.6) | 1,900 (3.8) |
| 90 | Constant rate | 223 (11.2) | 647 (10.8) | 1,900 (10.6) | 1,900 (3.8) |
|  | Logistic growth model | 226 (11.3) | 651 (10.8) | 1,900 (10.6) | 1,900 (3.8) |

**Fig 2.**
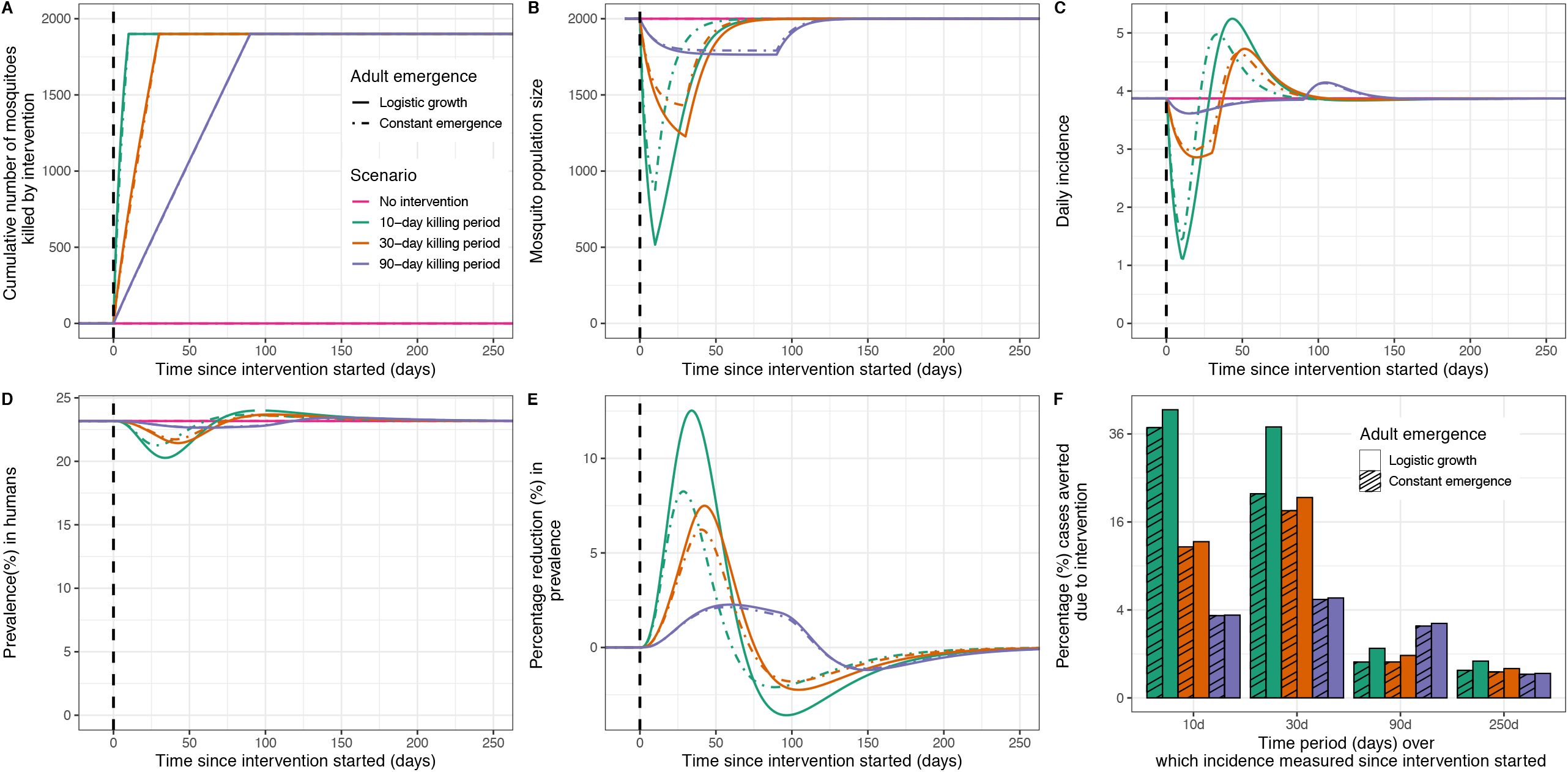
The theoretical impact of killing the same number of mosquitoes over different periods of time (10 days, 30 days and 90 days) on mosquito population and transmission dynamics of a generic vector borne disease, predicted by a simple SEIRS-SEI mathematical transmission model. A: The number of mosquitoes killed over the different time periods; B: the changes in mosquito population size due to killing mosquitoes; C: dynamics in daily disease incidence due to the intervention; D: dynamics of disease prevalence in humans and E: dynamics in the percentage reduction (%) in disease prevalence in humans in the intervention scenarios compared to a counterfactual scenario of no interventions (demonstrating how efficacy against prevalence would be measured in cross-sectional surveys). The vertical dashed line denotes the time at which the intervention started in the population. Scenarios are colour-coded: pink lines show the baseline scenario (no interventions), green show the 10-day killing period, orange show the 30-day killing period and purple the 90-day killing period. Adult emergence is assumed to be constant (dashed lines) or governed by a logistic growth model (solid line). Panel F: the efficacy of the different killing strategies, measured as the relative difference in the total number of cases for each killing strategy compared to a baseline scenario of no interventions. The efficacy of the different strategies is calculated over different time periods since the start of the intervention (10 days, 30 days and 90 days) and that by which prevalence and incidence have returned to endemic equilibrium (250 days since the start), which is used to report the overall efficacy of the different strategies. Solid bars indicate efficacy for model scenarios whereby adult emergence is assumed to be governed by a logistic growth model; hashed bars for those where adult emergence is assumed to be constant.

In the absence of interventions, malaria is endemic in these scenarios (thus dynamics are at an endemic equilibrium). The control intervention introduces changes to mosquito mortality and (and also to mosquito emergence if a logistic growth model is assumed to govern adult emergence), which drive the subsequent changes in daily incidence and malaria prevalence in humans (Fig 2A-D). In trial settings, the efficacy of vector control interventions can be measured in a number of ways, such as the percentage reduction in prevalence (from cross-sectional surveys) in the intervention arm compared to the control arm, or estimates of the percentage of cases averted (over a given time period) due to the intervention. Fig 2E shows model-predicted estimates of the percentage reduction in prevalence due to the interventions over time. Fig 2F shows the percentage of cases averted due to the interventions, measured at different time periods. Overall, the greatest reduction in incidence is for the intervention which kills mosquitoes in 10 days. However, for all time periods, there is a rebound in incidence and prevalence above the endemic equilibrium, due to the accumulation of people with lower population immunity in the period when mosquitoes are killed and lower force of infection (S1 FigC show how these dynamics contribute to fluctuations in the effective reproduction number, *R*_*e*_, over time). Therefore, the measurement of efficacy, here defined as either the difference in prevalence or the total cases averted (%) in each intervention group compared to a baseline scenario of no interventions, changes depending on how it is measured and when the measurement is taken (Fig 2F). Nevertheless, the difference caused by the assumptions on mosquito emergence on the overall efficacy of the intervention considered is modest (Fig 2F), and does not substantially impact the overall percentage of cases averted. When efficacy is measured at the point that all transmission dynamics have returned to endemic equilibrium, the cases averted for the 10-day, 30-day and 90-day interventions are very similar (for logistic growth: compared to a counterfactual scenario of no interventions, the 10-day killing intervention averts 0.7% of cases whereas the 90-day intervention averts 0.3% of cases; for constant emergence: 0.4% and 0.3% respectively.).

For trials conducting cross-sectional prevalence surveys at 28 days since the intervention has ceased, for a given assumption on adult mosquito emergence, a greater reduction in prevalence would be reported for a short-acting intervention compared to a longer-lasting intervention (for a logistic growth model, an approximately 12% reduction in all-age prevalence would be reported for the intervention lasting 10 days compared to a 2% reduction for the intervention lasting 90 days; for constant adult emergence, an approximately 6% reduction compared to a 2% reduction, respectively). The dynamics of the mosquito population will vary according to how killing adult mosquitoes impacts the number of mosquitoes emerging. For example, Fig 2E shows that if mosquito emergence remains constant, the peak in difference in prevalence between the arms happens 29 days after the 10-day long intervention ceased, whereas it was 34 days if assuming reducing mosquito numbers reduces mosquito emergence. If efficacy is measured later than planned, which is possible due to logistic constraints in field settings which may delay the timing of cross-sectional surveys, then the interventions may appear to have a negative effect due to the rebounds that occur (for example, assuming a logistic growth model, a negative reduction in prevalence (indicating a higher prevalence in the intervention arm compared to the control arm) would be detected for the 10-day intervention if the efficacy is measured approximately a month later than planned).

A sensitivity analysis of the same scenario in a setting with a higher level of endemicity (simulated using a higher vector-host ratio) was conducted. Here, we kill a total of 190,000 mosquitoes in a population with 200,000 mosquitoes at endemic equilibrium, which is proportional to the lower transmission setting where 1,900 mosquitoes were killed in a population with 2,000 mosquitoes at endemic equilibrium. In this higher transmission setting, it is similarly shown that if efficacy is measured over the period from the start of the intervention to when all transmission dynamics have returned to endemic equilibrium, the impact of the different killing strategies is also largely the same (S2 Fig). As mentioned, the estimates of efficacy in prevalence become negative if a loss of population immunity causes disease prevalence to bounce back higher than endemic endemic equilibrium once the transient impact of the vector control tool depreciating mosquito density has worn off. This emphasises the need to focus on the specific disease of interest and the importance of reliably quantifying population-level dynamics.

### Impact of synchronised and slow mass drug administration deployment of an ivermectin-like endectocide

An existing mathematical model of *P. falciparum* malaria transmission that has been used to explore the epidemiological impact of the MDA of an ivermectin-like endectocide [15, 28] was adapted to investigate how the time taken to complete MDA in a community impacts the predicted impact of the intervention. A community is defined as individuals living in a given geographical area who have the same broad risk of malaria (force of infection), though people within that community have different exposure to mosquito bites determined by their age, propensity to be bitten and use of control interventions. In all simulations, MDA of the ivermectin-like endectocide (3×300 *µ*g/kg dosage) is conducted monthly for 3 months, though the time to complete each round varies from a synchronised deployment (everyone due to receive the MDA takes the tablets on the same day), to over 20 days (slow deployment). We explore transmission in a perennial and seasonal setting (using the highly seasonal setting of Fatick, Senegal, as an example), with the MDA starting approximately 6 months into the year, shortly after mosquito densities rise in response to rainfall. Following the metrics recorded in ivermectin cRCTs, efficacy is measured in three ways; as the percentage of clinical cases averted in children under 5-years-old [20], from the start of the first MDA to 6 months later and the relative percentage difference in all-age prevalence at 4 weeks after the last MDA, between trial arms [29]. We additionally measure efficacy as the percentage of cases averted in children under 5-years old to 1 year later between trial arms.

Our analysis suggests that in both perennial and seasonal settings, the slow MDA deployment of the ivermectin-like endectocide (conducted over 20 days) has a greater epidemiological impact than the synchronised and 10-day deployments (Fig 3 & 4). These findings are observed regardless of how and when the efficacy measurement is taken. Similar results were observed in low transmission settings (S7 Fig - S9 Fig). Estimates are derived assuming a 70% coverage of the endectocide amongst eligible individuals and the error bars indicate the effect sizes at 50% and 90% coverage amongst eligible individuals. In the perennial setting (Fig 3), the 20-day and synchronised deployments avert a similar percentage of clinical cases in children under 5-years-old over one year since the intervention started (20-day deployment: 19% (17-21% at 50% and 90% coverage); synchronised strategy: 16% (14-17% at 50% and 90% coverage)). In the seasonal setting, across the range of coverages explored, the 20-day deployment averts more cases than the synchronised strategy (20-day strategy: 68% (60-74% at 50% and 90% coverage); synchronised strategy: 57% (50-60% at 50% and 90% coverage)).

**Fig 3.**
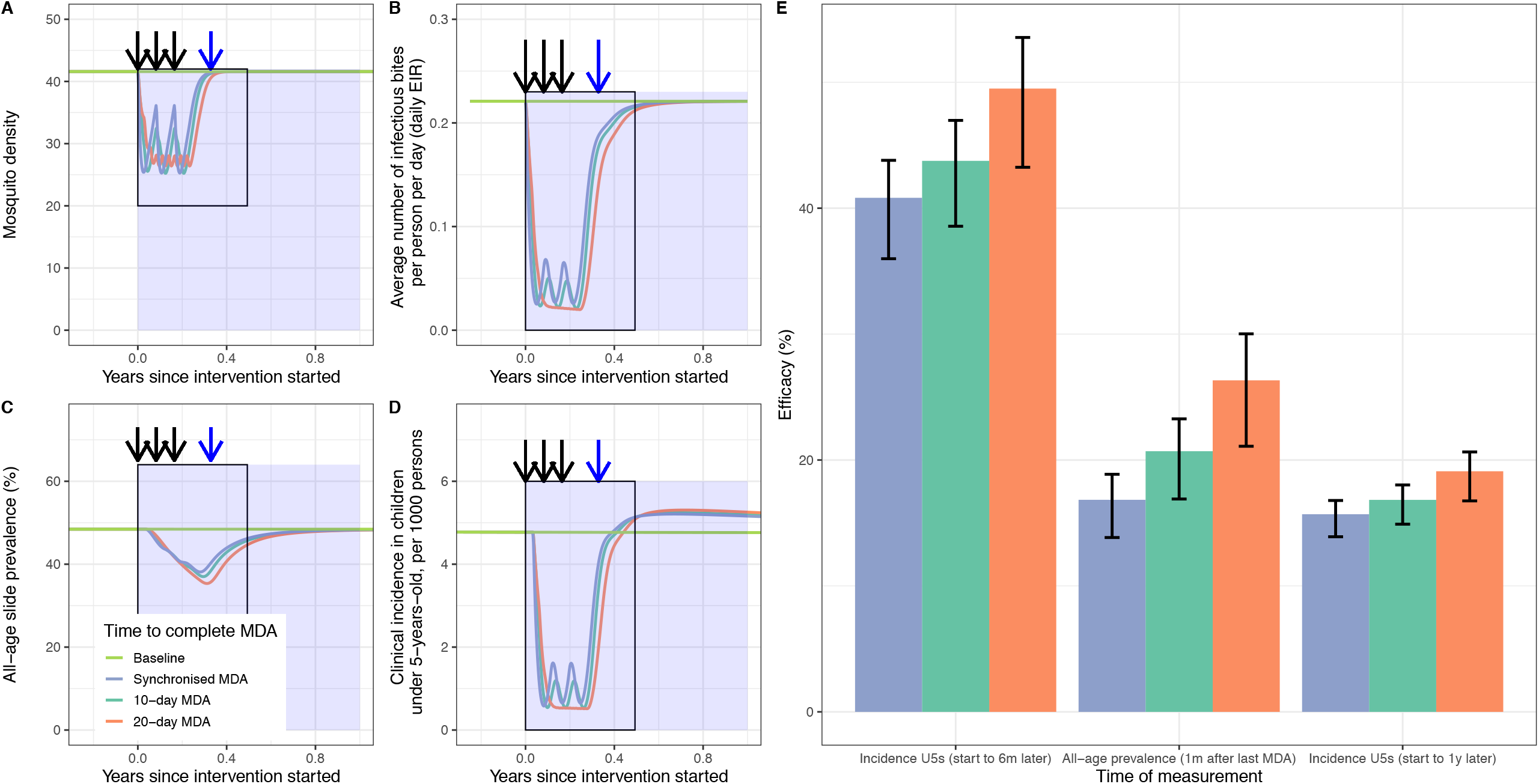
Transmission dynamics and efficacy of synchronised or slow MDA deployments of ivermectin-like endectocide in a high endemicity and perennial setting. Panels A-D show, respectively, mosquito density (vector-host ratio), daily entomological inoculation rate (EIR), malaria all-age slide prevalence (%) and clinical incidence of malaria in children under 5-years-old. Summary efficacy estimates are shown in Panel E. In Panels A-D, the light green line shows a scenario where no interventions are introduced, whilst the other colours show varying times for MDA deployment, be it synchronised, i.e. all individuals due to receive the MDA do so on the same day, (blue), 10 days (dark green) or 20 days (orange). MDAs take place once a month for 3 months (denoted by the 3 black arrows). The dark blue arrow in panel C shows when prevalence is measured (1 month after the last MDA, efficacy shown in middle set of bars of panel E). In panel D the black rectangular and the blue shaded area show, respectively, the 6-month and 12-month periods over which the cumulative number of clinical cases averted was calculated (the % difference from the no intervention arm shown as the other efficacy bars in panel E) . Panel E: The efficacy of synchronised and slow deployments, measured by calculating the relative reduction (%) in clinical incidence in children under 5-years-old compared to a baseline scenario of no interventions. The height of the bars corresponds to the efficacy derived from a simulated endectocide coverage of 70% amongst eligibles, with the vertical error bars correspond to a 50% (lower bound) or 90% (upper bound) simulated endectocide coverage.

We conducted a sensitivity analysis to explore the influence of starting the MDA later in the transmission season on the impact of the different distribution scenarios. In this instance, the first MDA started around the time of peak transmission, which is 60 days later than the start of the previously presented scenarios. The transmission model suggested that in this scenario, the slower distribution is less impactful (Fig 4E); the synchronised MDA averts approximately 27% of cases (24% - 29.4% at 50% and 90% coverage) whereas the slower 20-day MDA averts approximately 25% of cases (22.0% - 27.5% at 50% and 90% coverage). Nevertheless, the differences are modest and there is a considerably greater degree of variability in efficacy for a given deployment scenario due to endectocide coverage, than between scenarios.

**Fig 4.**
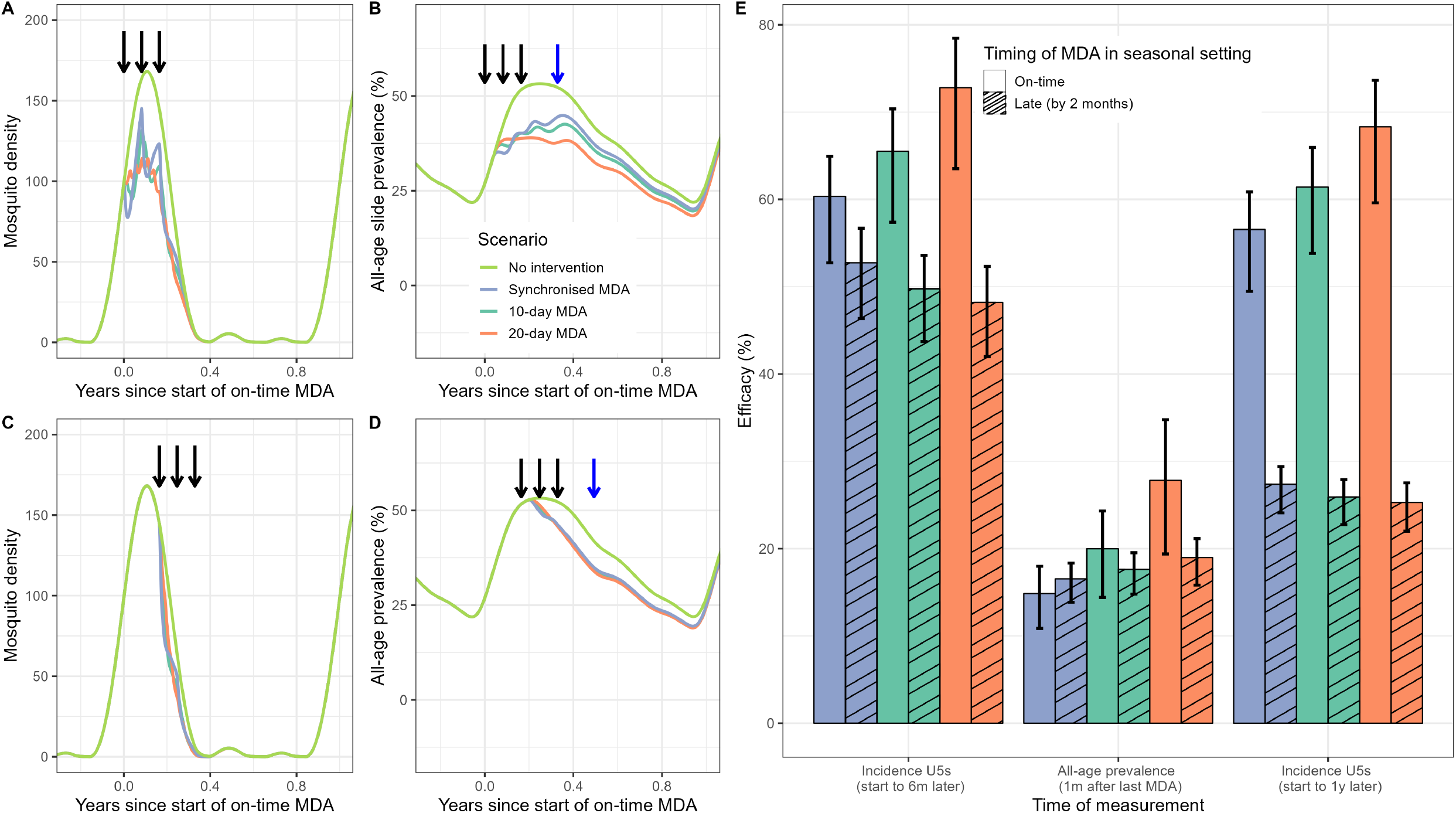
Transmission dynamics and efficacy of fast or slow MDA deployment of an ivermectin-like endectocide in a high endemicity and highly seasonal setting, where MDA deployment is either on-time, in relation to seasonality, or late, of which the latter can occur due to logistical constraints associated with MDA deployment. The late MDA campaigns are simulated to start 60 days later than the well-timed MDA, coinciding with the time of the peak in clinical incidence. Panels A and C show the dynamics in mosquito density (vector-host ratio), in relation to the start of the on-time MDA. Panels B and D show the dynamics in all-age prevalence, in relation to the start of the on-time MDA. In panels A-D, the light green line shows a scenario where no interventions are introduced, whilst the other colours show varying times for MDA deployment, be it the synchronised MDA (blue), 10-day MDA (dark green) or 20-day MDA (orange). MDAs take place once a month for 3 months (denoted by the 3 black arrows). Panels A and B show dynamics in the scenario where the MDA starts on-time, whereas panel C and D show dynamics where the MDA starts 60 days later than planned. The dark blue arrow in panels B and D show when prevalence is measured (1 month after the last MDA, efficacy shown in middle set of bars in panel E). Panel E: The efficacy of fast and slow deployment, measured by calculating the relative reduction (%) in clinical incidence in children under 5-years-old compared to a counterfactual scenario of no interventions, measured from the start of the MDA to 6 months later, as the relative reduction in prevalence (%) between the counterfactual scenario and intervention scenario at 4 weeks after the last MDA and the relative reduction (%) in clinical incidence in children under 5-years-old compared to a counterfactual scenario of no interventions. Solid bars show the efficacy of the on-time MDA, and the hashed bars show the efficacy of the MDA that is 2 months late to start. The height of the bars corresponds to the efficacy derived from a simulated endectocide coverage of 70% amongst the eligible population, with the vertical error bars correspond to a 50% (lower bound) or 90% (upper bound) simulated endectocide coverage amongst eligibles.

## Discussion

A simple vector-borne disease model was used to explore whether the time taken to kill mosquitoes influences the epidemiological impact of interventions. On average, the results suggest minimal difference in disease incidence, regardless of whether the same number of mosquitoes are killed over 10 days, 30 days or 90 days. Though entirely theoretical, this result could have implications for future insecticidal product development, which should concentrate on maximising the number of vectors killed rather than achieving this within a particular time-frame. Nevertheless, even simple models can generate complex dynamics when pulsed interventions interact with seasonality of transmission and wider infectious disease dynamics. This highlights the need for bespoke transmission dynamics models in order to understand how efficacy measures can change depending on how and when disease burden is measured. The work again highlights how non-linear relationships between mosquito density and disease transmission dynamics [31] need to be considered when evaluating the impact of vector control interventions which kill mosquitoes.

The simple models also illustrate how the density-dependent processes which regulate the number of vectors in the population can influence disease dynamics and the observed effectiveness of interventions. If it is assumed that adult emergence is governed by a logistic growth model, then the impact of the interventions are larger as the adult emergence rate is heavily influenced by the mosquito population size at a given time point (*M*_*t*_), leading to a reduced emergence rate when the intervention is applied, compared to if constant adult emergence is assumed (Fig 2.B). In the latter scenario, mosquitoes emerge at a constant rate regardless of the number of mosquitoes that are killed. This could occur in real-world settings, if for example, the mosquitoes that are killed by a control interventions in a community emerge from breeding sites that are far away from the community and thus unaffected by the intervention. The intervention is consistently more efficacious if a logistic growth model is assumed to govern adult mosquito emergence (Fig 2F). The implications of reductions and rebounds in transmission dynamics due to disease control interventions have been explored in ecology for other infectious diseases, such as rodent control to prevent Lassa virus [32]. These authors combined a field study with mathematical modelling and found that as rodent populations recover so quickly after application of annual rodenticide, continuous control is required to minimise Lassa virus spillover. When extrapolating our findings to other malaria vector control interventions, it is important to consider that this modelling framework does not have a spatial component (i.e. it is not spatially explicit) and assumes a constant additional mortality rate (due to the intervention) over time. These factors may not hold in real-world settings where spatial segregation of mosquito populations does occur and that maintaining intervention efficacy and coverage over time is challenging due to the waning effects of insecticides and human factors which lead to reduced coverage [8, 33]. The simple transmission model used in this study can therefore be used to theoretically explore the implications of killing the same number of mosquitoes over different time periods, however it is unlikely that this exact scenario would occur in real-world settings.

In recent years, a number of cRCTs evaluating the impact of ivermectin MDA against malaria transmission took place in Africa. The trial sites included settings in Burkina Faso, Guinea-Bissau, The Gambia, Mozambique and Kenya [20, 29, 30, 34]. These trials, with the exception of the study conducted in Mozambique, reported no evidence of the epidemiological impact of ivermectin on malaria transmission. Transmission dynamics mathematical models are increasingly being used to support the trial design process as they provide a framework to enable evidence-based estimation of effect size and other metrics of epidemiological importance used to support power calculations [35]. Just as it is important to understand why the trial did not generate the expected result, it is also necessary to explore why empirical evidence differs from model predictions. It is necessary to differentiate whether this is either a structural issue with the model (i.e. the model is unable to reliably translate an entomological impact into changes in the epidemiology of malaria), or whether the parameterisation of the intervention in the model did not capture the effect of the drug on mosquito populations in the real world. The ivermectin model used to guide the trials [28] makes sets of entomological and epidemiological assumptions based on data from an early phase II clinical trial (the IVERMAL trial) [13]. The model assumes the same relative increase in mortality and duration of killing for laboratory-reared *An. gambiae sensu stricto* mosquitoes (as observed in the IVERMAL trial) and wild-caught mosquitoes. This assumption may not hold, as biological differences between wild and laboratory-reared mosquitoes are known to exist, for example, the life expectancy of former is generally considerably longer than the latter [36]. As new data emerges from the recently completed trials, it will become possible to assess whether these assumptions were indeed valid and could have driven the lower than expected epidemiological impact. The model presented here could be used to assess the sensitivity of the trial results to assumptions on both the mosquito-killing efficacy of ivermectin and the time to complete MDA campaigns. For example, emerging data from the BOHEMIA trial suggests that the mosquito-killing effect of the 3×300 *µ*g/kg dosage (estimated from field data) is lower than previously suggested by the IVERMAL trial [13, 37], and recent modelling work has suggested that this would result in a lower epidemiological impact of an ivermectin-like endectocide [38]. The differences between ivermectin-induced mortality of wild vs. laboratory-reared mosquitoes is one possible explanation, amongst others such as heterogeneities in susceptibility to ivermectin amongst *Anopheles* species and insecticide-resistance phenotypes [39], of the null results observed in recently completed cRCTs.

Models can be used to explore whether programmatic details of the trial could have contributed to the hiding of an epidemiological impact when one was indeed present. Various hypotheses have been proposed for the different results in the different ivermectin cRCTs, making it unclear whether the intervention does not work or the cRCTs, for several possible reasons, were adequately powered to detect an impact. For example, the RIMDAMAL II trial in Burkina Faso (ClinicalTrials.gov: NCT03967054) found a null result, and it was suggested that the mass distribution of the highly effective Interceptor-G2 ITNs, shortly after the ivermectin MDA campaigns, reduced incidence in both arms, leaving the trial underpowered to detect a difference in impact [40]. The MATAMAL trial also found a null result, and it has been hypothesised that one reason for this (amongst others) was related to the biting behaviours of *Anopheles gambiae sensu lato* mosquitoes on the islands in which the trial took place, given evidence of zoophagic behaviours for coastal vectors on mainland Guinea-Bissau [18, 41]. The modelling used to guide and power these trials did not consider such differences in biting behaviours due to a lack of data, at the time, from the trial sites, potentially influencing the model-predicted effect sizes and power calculations. As mentioned, another hypothesis is that the mathematical models used to support trial design assumed that all individuals due to receive the MDA did so at the same time [14, 15], whereas in reality, on average, MDA campaigns took at least 9 days to complete and even 20 days in a setting where a tropical cyclone limited the logistical feasibility of conducting the MDA quickly [19, 21]. Overall, it is likely that the initial modelling work which guided trials oversimplified many real-world complexities that could have limited the impact of the endectocide in trial settings. Nevertheless, our modelling results suggest that the efficacy of the MDA of an ivermectin-like endectocide is greater when implemented slowly, in both perennial and seasonal settings, providing evidence that it was not the slow MDA programme alone which drove the differences in epidemiological results. These findings are consistent with previous modelling work exploring the impact of the time taken to complete the MDA of antimalarial drugs, which suggested that in low mobility populations, slower MDA campaigns of antimalarial drugs have a greater impact than fast MDAs, however in this instance, this is due to the slower build-up of resistant infections [42]. Other factors, for example, the potential influence of other vector control interventions on the relative impact of ivermectin (through shortening mosquito lifespan and affecting the time between bloodmeals on humans [25]) and the proportion of the population that were eligible for the intervention or refused it, may have affected the real-world impact of ivermectin as an endectocide for malaria control and could warrant further model exploration.

Our sensitivity analysis on the timing of the endectocide MDA in relation to seasonality suggests that if the MDA starts too late in the season, impact is minimised. In the Mozambique site of the BOHEMIA trial, the MDA started later than planned due to the severe logistical constraints imposed by cyclone Gombe, making it challenging for field teams to commence the MDA [19]. Timing the use of short-acting interventions appropriately will likely be difficult, especially when climatic changes and natural disasters can drive changes in rainfall and malaria seasonality that are difficult to predict [43]. Seasonality in mosquito population dynamics is likely more complex than captured in the transmission model used in this study, and can be characterised by further peaks and troughs [44]. Subsequently, the impact of the three consecutive doses (3×300*µ*g/kg) of an ivermectin-like endectocide efficacy could be less impactful if deployments coincide with troughs in mosquito density. Spacing out these three consecutive doses could therefore potentially be more beneficial. Atypical rainfall can also change malaria endemicity, which was observed after the cyclone in Mozambique [45], which could compromise power calculations as unexpected climatic events are not traditionally considered in the transmission modelling used to guide trials. Longer-lasting formulations of endectocides, e.g. those with longer half-life than ivermectin, such as the drug, nitisinone [46], may be more robust to the impacts of seasonality on epidemiological impact, although thus far the safety profile of nitisinone in healthy individuals is unknown and further modelling work would need to be conducted to test this endectocide against a range of potential seasonalities of transmission. Overall, in this changing landscape of malaria disease ecology, it is important that vector control interventions are robust to variability in seasonality, and identifying products that are both safe in humans, with minimal ecological and environmental impact and with a sufficiently long half-life will be essential to the design of endectocides in the future.

There are limitations on the generalisability of our findings. Firstly, the impact of other vector control interventions on malaria transmission and the relative impact of endectocides has not been considered. For example, ITNs and IRS may influence the effectiveness of endectocides by shortening the mosquito lifespan or the time between bloodmeals on humans [25]. However, it is unlikely that these would change within the time periods of the MDA (10 days or 20 days). Our predicted magnitudes of the impact of ivermectin-like endectocides in perennial and seasonal settings may also be exaggerated by the model assumptions on temporal trends in mosquito density, which are likely more complex in wild mosquito population and are also characterised by differences in population age structure [44]. We have also not considered correlations between heterogeneity in mosquito biting and endectocide coverage, for example, that certain populations may be more at risk of being bitten by mosquitoes [47] and that these same populations may be less likely to be treated with the ivermectin-like endectocide (for example, due to being hard-to-reach). This could be taken into account in future work by incorporating additional model compartments with different levels of biting risk and endectocide coverage. Our slower endectocide deployment scenarios are simulated using evenly interspaced distributions (S5 FigA), and this is likely different to how the intervention is implemented on the ground. For example, in the MATAMAL trial in Guinea-Bissau, 80% of the MDA was completed within 10 days (on average) in each arm, however, there was a mop-up period to search for absentees which increased the overall time to complete the MDAs to 24 days [18]. Future modelling work using local data for model parameterisation and fitting may help to evaluate the use-case of endectocides in particular settings.

Overall, our theoretical modelling exercise evaluates the extent to which the time taken to kill mosquitoes influences the epidemiological impact of vector control interventions. Using a simple model, we demonstrate that differences in the time taken to kill a target number of vectors does not lead to detectable differences in epidemiological impact. Our more complex malaria model of ivermectin-like endectocide MDA supports these results. Improving the realism of vector-borne disease models can enable more reliable predictions of the epidemiological impact of potential control interventions. Our work suggests that for a given endectocide coverage, slower MDA campaigns lead to a greater number of clinical *P. falciparum* cases averted in children under 5-years-old in perennial settings and in seasonal settings (if the intervention starts on time), compared to synchronised distributions, as a greater proportion of the treated population have a lethal dose of the ivermectin-like endectocide at any given time, leading to more mosquitoes being killed (S5 FigC). However, the model suggests that if the MDA starts too late in relation to seasonality, the impact of the intervention is substantially minimised. This work accentuates the variability of the epidemiological impact of ivermectin-like endectocides for malaria control, depending on the time taken to complete the MDA and when the MDA is conducted, which can vary due to unpredictable factors associated with operational challenges in achieving the desired coverage and sudden seasonal changes or natural disasters which influence mosquito population dynamics and subsequently, malaria transmission dynamics in a non-linear fashion.

## Supporting Information

**S1 Text. Supplementary Methods**.

**S1 Fig. Dynamics in infectious vectors (%), susceptible humans (%) and the effective reproduction number**, *R*_*e*_(*t*) **in the low endemicity setting (there are 2 vectors per host)**.

**S2 Fig. The theoretical impact of killing the same number of mosquitoes over different periods of time (10 days, 30 days and 90 days) on mosquito population and *Plasmodium* falciparum malaria transmission dynamics in a high endemicity setting (there are 200 vectors per host), predicted by a simple SEIRS-SEI mathematical transmission model**.

**S3 Fig. Schematic of the modelling structure of the human malaria model (adapted from Supporting Information Material 1.1 [15]**.

**S4 Fig. Daily hazard ratios for mosquito death the 3×300***µ***/kg dosage of ivermectin, from a phase II trial [27]**.

**S5 Fig. Population-level hazard ratios and time-varying proportion of people with a toxic dose of ivermectin, used for simulating prolonged and synchronised distributions of ivermectin MDA, taking place monthly for three months**.

**S6 Fig. Transmission dynamics (daily EIR per person and clinical incidence of malaria in children under 5-years-old in a high endemicity and highly seasonal setting, where MDA deployment of an ivermectin-like endectocide is either on-time, in relation to seasonality, or late, of which the latter can occur due to logistical constraints**.

**S7 Fig. Transmission dynamics and efficacy of synchronised or slow MDA deployments of ivermectin-like endectocide in a low endemicity and perennial setting**.

**S8 Fig. Transmission dynamics and efficacy of synchronised or slow MDA deployments of ivermectin-like endectocide in a low endemicity and highly seasonal setting, where the timing of the MDA is deemed on-time in relation to seasonality in mosquito abundance**.

**S9 Fig. Transmission dynamics and efficacy of synchronised or slow MDA deployments of ivermectin-like endectocide in a low endemicity and highly seasonal setting, where the timing of the MDA is late in relation to seasonality in mosquito abundance**.

**S1 Table. Values used to parameterise simple malaria transmission model**. Mean values are shown for all parameter values expressed as periods.

**S2 Table. Summary of the number (and percentage) of mosquitoes killed by the interventions with different mosquito-killing periods (10 days, 30 days or 90 days), as predicted by a generic SEIRS-SEI model of vector-borne disease transmission in which there are approximately 200 mosquitoes per person**. The percentage of mosquitoes killed by the intervention was calculated by tracking the total number of mosquitoes killed by the intervention (the D compartment) compared to total number of mosquitoes to have emerged into the population (in the counterfactual no-intervention scenario) from the start of each mosquito-killing period to 10 days, 30 days, 90 days and 250 days later (All dynamics have returned to equilibrium by 250 days after the start of the mosquito-killing started). By each of these time points, 200,000, 600,000, 1,800,000 and 5,000,000 mosquitoes have emerged into the population, respectively.

## Acknowledgments

We thank the RESIDE team at the MRC Centre for Global Infectious Disease Analysis for useful software support, required for undertaking this project. We also thank the Ivermectin for Malaria Elimination Network (IVERMEN) for useful discussions on the logistics of implementing ivermectin MDA campaigns.

