## Supporting Information for "Modelling the influence of the duration of effectiveness and time taken to implement vector control interventions for vector-borne diseases"

### 1 Supporting Information

#### 1.1 Simple SEIRS-SEI model

As described in the main text, a simple deterministic and mechanistic malaria transmission model was used to explore the influence of killing a target number of adult female mosquitoes (D) over three different time periods (10 days, 30 days or 90 days) on mosquito population dynamics, malaria transmission dynamics and epidemiological impact. The model was implemented using the *odin* R package [1].

The differential equations governing transmission dynamics in humans are shown in Supporting Information equations 1-4. The number of new human malaria cases (incidence) is tracked by the rate at which humans enter the E compartment, as shown in Supporting Information Equation 5. Model parameters are defined in Table refsm-tab:simple-model-ref. Given the total human population, $N = S_h + E_h + I_h + R_h$, the human model is:

$$\frac{dS_h}{dt} = \mu_h N + \omega_h R_h - \left(\beta_{hv}\frac{I_v}{N} + \mu_h\right) S_h \tag{1}$$

$$\frac{dE_h}{dt} = \beta_{hv} S_h \frac{I_v}{N} - (\sigma_h + \mu_h) E_h \tag{2}$$

$$\frac{dI_h}{dt} = \sigma_h E_h - (\gamma_h + \mu_h) I_h \tag{3}$$

$$\frac{dR_h}{dt} = \gamma_h I_h - (\omega_h + \mu_h) R_h \tag{4}$$

$$\frac{dC_h}{dt} = \beta_{hv} S_h \frac{I_v}{N} \tag{5}$$

Supporting Information Equations 6 - 8 show the differential equations for the simple SEI framework for modelling malaria infection in mosquitoes. When the intervention is on (from $\tau$, when the intervention begins, over $\Delta t$, the duration of the intervention), an additional mortality rate ($\psi_v$) is exerted onto the mosquito population. The total number of adult female mosquitoes killed by the intervention is tracked using a D compartment (Supporting Information Equation 9). Model parameters are defined in Table refsm-tab:simple-model-ref. Two variants ($i \in \{1, 2\}$) of the model are considered whereby either a constant emergence rate ($i = 1$) or a logistic growth model ($i = 2$) for the adult female mosquito population is assumed. Given the total adult female mosquito population, $M = S_v + E_v + I_v$, the mosquito model is:

$$\frac{dS_v}{dt} = \phi_{v,i} - \left(\beta_{vh}\frac{I_h}{N} + \mu_v + \psi_v(t)\right) S_v \tag{6}$$

$$\frac{dE_v}{dt} = \beta_{vh} S_v \frac{I_h}{N} - \left(\sigma_v + \mu_v + \psi_v(t)\right) E_v \tag{7}$$

$$\frac{dI_v}{dt} = \sigma_v E_v - \left(\mu_v + \psi_v(t)\right) I_v \tag{8}$$

$$\frac{dD_v}{dt} = \psi_v(t) M \tag{9}$$

Where $r$, the instrinsic growth rate, is assumed to be equal to $\mu_v$:

$$\psi_v(t) = \begin{cases} \psi_v & \text{if } \tau \leq t \leq \tau + \Delta t \\ 0 & \text{otherwise} \end{cases}$$

$$\phi_i = \begin{cases} \mu_v M_0 & \text{if } i = 1 \\ \mu_v M + rM\left(1 - \frac{M}{M_0}\right) & \text{if } i = 2 \end{cases}$$

Noting that under the definition of $\phi_{v,i=2}$, a classical logistic growth model is recovered for the overall adult female mosquito population in the absence of the intervention:

$$\frac{dM}{dt} = \begin{cases} \mu_v M_0 - \mu_v M - \psi_v(t) M & \text{if } i = 1 \\ \mu_v M + rM\left(1 - \frac{M}{M_0}\right) - \psi_v(t) M & \text{if } i = 2 \end{cases} \tag{10}$$

Here, the basic reproduction number, $\mathcal{R}_0$, is defined as the expected number of secondary human infections generated by a single primary human infection in an otherwise susceptible human and adult female mosquito population, and was derived from the ODE system (Supporting Information equations 1-4 and refsm-eq:ento-Sv-8) using a next-generation matrix method. The ODE system was linearised at the disease-free equilibrium ($S_h = N_0$, $S_v = M_0$) and the transmission, $F$, and transition, $V$, matrices were then

**Table S1. Values used to parameterise simple malaria transmission model.** Mean values are shown for all parameter values expressed as periods.

| **Model parameter** | **Definition** | **Value or functional form** | **Reference** |
|---|---|---|---|
| $N_0$ | Initial human population size | 1000 | - |
| $\mu_h^{-1}$ | Human life expectancy | 70 years | - |
| $\sigma_h^{-1}$ | Intrinsic incubation period | 12 days | [2] |
| $\gamma_h^{-1}$ | Human infectious period | 60 days | [3] |
| $\omega_h^{-1}$ | Human duration of protective immunity | 180 days | [4] |
| $a^{-1}$ | Mosquito gonotrophic cycle | 2 days | [5] |
| $b_{hv}$ | Probability of vector to human transmission per bite | 0.6 | [6] |
| $\beta_{hv}$ | Effective daily vector to human transmission rate | $= ab_{hv}$ | - |
| $M_0$ | Initial mosquito population size | 2000 | - |
| $\mu_v^{-1}$ | Mosquito lifespan | 10 days | [7] |
| $\sigma_v^{-1}$ | Extrinsic incubation period | 10 days | [8] |
| $m$ | Ratio of adult female mosquitoes to humans | $= M/N$ | - |
| $b_{vh}$ | Probability of human to vector transmission per bite | 0.6 | assumed to equal $b_{hv}$ |
| $\beta_{vh}/m$ | Effective daily human to vector transmission rate | $= ab_{vh}/m$ | - |

computed. The dominant eigenvalue of the next generation matrix, $FV^{-1}$, was then derived and squared to account for two epidemiological generations elapsing in the transmission cycle (human-to-vector and vector-to-human), yielding an expression for the basic reproduction number in the absence of any intervention:

$$\mathcal{R}_0 = \frac{\beta_{hv}\beta_{vh}\sigma_h\sigma_v m}{(\sigma_h + \mu_h)(\gamma_h + \mu_h)(\sigma_v + \mu_v)\mu_v} \tag{11}$$

The same approach was then followed to derive an expression for the controlled reproduction number ($\mathcal{R}_c$) in the presence of the intervention for fully susceptible adult female mosquito and human populations:

$$\mathcal{R}_c = \left(\frac{(\sigma_v + \mu_v)\mu_v}{(\sigma_v + \mu_v + \psi_v)(\mu_v + \psi_v)}\right)\mathcal{R}_0 \tag{12}$$

The instantaneous potential reproduction number over time ($\mathcal{R}$) for fully susceptible populations is therefore:

$$\mathcal{R}(t) = \begin{cases} \mathcal{R}_c & \text{if } \tau \leq t \leq \tau + \Delta t \\ \mathcal{R}_0 & \text{otherwise} \end{cases} \tag{13}$$

While the instantaneous effective reproduction number ($\mathcal{R}_e$) in the presence of transmission can then be inferred from:

$$\mathcal{R}_e(t) = \left(\frac{S_h(t)}{N}\frac{S_v(t)}{M(t)}\right)\mathcal{R}(t) \tag{14}$$

The endemic equilibrium ($S_h^*$, $E_h^*$, $I_h^*$, $R_h^*$, $S_v^*$, $E_v^*$, $I_v^*$) of the ODE system was then calculated by setting Supporting Information equations 1-4 and 6-8 equal to zero and solving for the non-trivial stable steady state in the absence of the intervention when $\mathcal{R}_0 > 1$:

$$S_h^* = N_0 - (E_h^* + I_h^* + R_h^*) \tag{15}$$

$$E_h^* = \left(\frac{\gamma_h + \mu_h}{\sigma_h}\right) I_h^* \tag{16}$$

$$I_h^* = \frac{N_0 \mu_v (\mathcal{R}_0 - 1)}{K} \tag{17}$$

$$R_h^* = \left(\frac{\gamma_h}{\omega_h + \mu_h}\right) I_h^* \tag{18}$$

$$S_v^* = M_0 - (E_v^* + I_v^*) \tag{19}$$

$$E_v^* = \left(\frac{\mu_v}{\sigma_v}\right) I_v^* \tag{20}$$

$$I_v^* = \frac{\beta_{vh} \sigma_v M_0 I_h^*}{(\sigma_v + \mu_v)(\mu_v N_0 + \beta_{vh} I_h^*)} \tag{21}$$

Where:

$$K = \mathcal{R}_0 \mu_v \left(1 + \frac{\gamma_h + \mu_h}{\sigma_h} + \frac{\gamma_h}{\omega_h + \mu_h}\right) + \beta_{vh}$$

Since malaria is endemic in the modelled setting, the endemic equibrium conditions were used to set the initial conditions for the differential equations.

To investigate whether interventions that are in place for different durations, but kill the same number of mosquitoes, have different epidemiological impacts, although not modelled, we first consider the case of equal natural birth and death rates in the mosquito population:

$$\phi_0 = \mu_v M \implies \frac{dM}{dt} = -\psi_v(t)M \implies M(t) = M_0 e^{-\psi_{v0}(t)} \iff \tau \leq t \leq t + \Delta t \tag{22}$$

where $\psi_{v0}$ is the additional mortality rate due to the intervention under the assumption of equal natural mosquito birth and death rates. Since $\frac{dD_v}{dt} = \psi_v(t)M$, the total number of adult female mosquitoes killed over the duration of an intervention ($\Delta D_0$) can then be calculated from:

$$\Delta D_0(\psi_{v0}, \Delta t; M_0) = \int_0^{\Delta t} \psi_{v0} M(s)\ ds = \psi_{v0} M_0 \int_0^{\Delta t} e^{-\psi_{v0} s}\ ds = M_0 \left(1 - e^{-\psi_{v0}\Delta t}\right) \tag{23}$$

given $s = t - \tau$ is the time since the intervention is introduced. Therefore, when mosquito birth and death rates are equal, for a given number of mosquitoes, $\Delta D_0$, to be killed by the intervention over its entire duration, $\Delta t$, we require the additional mortality rate due to the intervention to be:

$$\psi_{v0} = -\frac{1}{\Delta t} \log\left(1 - \frac{\Delta D_0}{M_0}\right) \iff \Delta D_0 < M_0 \tag{24}$$

We then consider the first modelled case for a constant emergence of adult female mosquitoes. In an analogous manner to Supporting Information Equation 23, under the assumption of a constant emergence rate, the number of mosquitoes killed over the intervention, $\Delta D_1$, can be calculated from:

$$\Delta D_1(\psi_{v,i=1}, \Delta t; M_0, \mu_v) = \frac{\psi_{v,i=1} M_0}{\mu_v + \psi_{v,i=1}} \left[\mu_v \Delta t + \frac{\psi_{v,i=1}}{\mu_v + \psi_{v,i=1}} \left( 1 - e^{-(\mu_v + \psi_{v,i=1})\Delta t}\right)\right] \tag{25}$$

Following the same approach again, the total number of mosquitoes killed by the intervention under the logistic growth model, $\Delta D_2$, can be shown to be:

$$\Delta D_2(\psi_{v,i=2}, \Delta t; M_0, \mu_v) = \begin{cases} M_0 \log(1 + \mu_v \Delta t) & \text{if } \psi_{v2} = \mu_v \\ \frac{\psi_{v,i=2} M_0}{\mu_v} \log\left(\frac{\mu_v e^{(\mu_v - \psi_{v,i=2})\Delta t} - \psi_{v,i=2}}{\mu_v - \psi_{v,i=2}}\right) & \text{if } \psi_{v,i=2} \neq \mu_v \end{cases} \tag{26}$$

Since neither the intervention mortality rate under the assumption of constant emergence ($\psi_{v,i=1}$) or logistic growth ($\psi_{v,i=2}$) can be expressed in a closed form solution by rearrangement of Supporting Information

Equations 25 or 26, to calculate the intervention mortality rate required to kill a desired, $\Delta D_{\text{target}}$, number of mosquitoes, we sought the value $\psi_{v=i}$ that would minimise the following objective function:

$$f(\psi_{v=i}; \Delta D_i, \Delta t, M_0, \mu_v) = [\Delta D_{\text{target}} - \Delta D_i(\psi_{v-i}, \Delta t; M_0, \mu_v)]^2 \quad (27)$$

This was conducted using the *optim* function in *R* with the *L-BFGS-B* method [9]; the analytical solution for the intervention mortality rate under equal birth and death rates ($\psi_{v0}$ from Supporting Information Equation 24) was used as an initial parameter guess for $\psi_{v=i}$.

The dynamics in infectious vectors and susceptible humans is shown in S1 Fig. The sensitivity analysis on the influence of the vector-host ratio on the impact on killing strategies with different durations of killing is shown in S2 Fig.

At the higher vector-host ratio (200 mosquitoes per person), the percentage of mosquitoes killed by each of the mosquito-killing period implementations is shown in Supporting Information Table S2.

| Mosquito-killing period (days) | Adult mosquito emergence assumption | N (%) of mosquitoes killed by intervention since it started | | | |
|---|---|---|---|---|---|
| | | 10 days | 30 days | 90 days | 250 days |
| 10 | Constant rate | 190,000 (8.20) | 190,000 (6.99) | 190,000 (4.85) | 190,000 (2.67) |
| | Logistic growth model | 190,000 (8.40) | 190,000 (7.45) | 190,000 (5.08) | 190,000 (2.74) |
| 30 | Constant rate | 70,478 (3.04) | 190,000 (7.00) | 190,000 (4.85) | 190,000 (2.67) |
| | Logistic growth model | 74,178 (3.21) | 190,000 (7.13) | 190,000 (4.94) | 190,000 (2.70) |
| 90 | Constant rate | 22,347 (0.97) | 64,747 (2.38) | 190,000 (4.85) | 190,000 (2.67) |
| | Logistic growth model | 22,570 (0.98) | 65,136 (2.40) | 190,000 (4.88) | 190,000 (2.68) |

**Table S2. Summary of the number (and percentage) of mosquitoes killed by the interventions with different mosquito-killing periods (10 days, 30 days or 90 days), as predicted by a generic SEIRS-SEI model of vector-borne disease transmission in which there are approximately 200 mosquitoes per person**. The percentage of mosquitoes killed by the intervention was calculated by tracking the total number of mosquitoes killed by the intervention (the D compartment) compared to total number of mosquitoes to have emerged into the population (in the counterfactual no-intervention scenario) from the start of each mosquito-killing period to 10 days, 30 days, 90 days and 250 days later (All dynamics have returned to equilibrium by 250 days after the start of the mosquito-killing started). By each of these time points, 200,000, 600,000, 1,800,000 and 5,000,000 mosquitoes have emerged into the population, respectively.

**Fig S1. Dynamics in infectious vectors (%), susceptible humans (%) and the effective reproduction number, $\mathcal{R}_e(t)$ in the low endemicity setting (there are 2 vectors per host)**. The model used is the simple SEIRS-SEI model of a generic vector-borne disease, parameterised here for malaria (other dynamics shown in Figure reffig2). In the model, the target number of mosquitoes (n = 1900) were killed over 10 days (green line), 30 days (orange line) or 90 days (purple line). The counterfactual scenario of no interventions (baseline) is shown by the pink line. Solid lines indicate that the model parameterisation for adult emergence is assumed to be constant, whereas for the dashed line it is assumed to be governed by a logistic growth model. Panel A shows dynamics in infectious vectors (%); B: dynamics in susceptible humans (%) and C: dynamics in $\mathcal{R}_e(t)$.

**Fig S2. The theoretical impact of killing the same number of mosquitoes over different periods of time (10 days, 30 days and 90 days) on mosquito population and *Plasmodium* falciparum malaria transmission dynamics in a high endemicity setting (there are 200 vectors per host), predicted by a simple SEIRS-SEI mathematical transmission model.** A: The number of mosquitoes killed over the different time periods; B: the changes in mosquito population size due to killing mosquitoes; C: dynamics in daily *P. falciparum* incidence due to the intervention and D: dynamics of *P. falciparum* prevalence in humans and E: dynamics in the effective reproduction number ($R_t$) over time. The vertical dashed line denotes the time at which the intervention started in the population. Scenarios are colour-coded: pink lines show the baseline scenario (no interventions), green show the 10-day killing period, orange show the 30-day killind period and purple the 90-day killing period. Adult emergence is assumed to be constant (solid lines) or governed by a logistic growth model (dashed line). Panel E: the efficacy of the different killing strategies, measured as the relative difference in the total number of cases for each killing strategy compared to a baseline scenario of no interventions. The efficacy of the different strategies is calculated over different time periods since the start of the intervention (10 days, 30 days and 90 days) and that by which prevalence and incidence have returned to equilibrium (200 days since the start), which is used to report the overall efficacy of the different strategies. Solid bars indicate efficacy for model scenarios whereby adult emergence is assumed to be governed by a logistic growth model; hashed bars for those where adult emergence is assumed to be constant.

#### 1.2 Malaria transmission with ivermectin mass drug administration

The mathematical modelling approach used to simulate the impact mass drug administration (MDA) of the endectocide, ivermectin, was previously implemented [10] in an existing malaria transmission model [3, 11] and can be accessed using the *ivRmectin* R package [12]. In the present study, we adapted this model to incorporate the effects of staggered distributions of ivermectin on mosquito mortality and subsequently, malaria transmmission dynamics.

The human component of this malaria model consists of 6 states of infection/disease. Each state is separated into different biting heterogeneity groups (capturing differences in the risk of exposure to infectious bites), divided into 20 age groups and into intervention groups, depending on the usage of the intervention by the population. Naturally acquired immunity parameters were previously fitted to capture age-specific epidemiological trends in different settings [13]. Once an individual has received an infectious bite, the probability of developing symptoms is dependent on immunity, which in turn is determined by the age and level of exposure in the population. The force of infection from mosquitoes to humans is dependent on, amongst other factors, the number of infectious vectors. The human model is illustrated in S3 Fig.

**Fig S3. Schematic of the modelling structure of the human malaria model (adapted from Supporting Information Material 1.1 [10]**. $FOI_{hv}$ denotes the force of infection on humans from vectors.

Malaria infection in mosquitoes is modelled in three states; susceptible ($S_v$), pre-infectious ($E_v$) and infectious ($I_v$) mosquitoes. Mosquitoes become infected upon biting an infectious person, according to the force of infection from humans to vectors ($\Lambda^v$, see Supporting Information Equation 29).

Mosquitoes surviving the extrinsic incubation period (EIP) become infectious and are able to transmit malaria to humans upon biting. Mosquitoes search for bloodmeals for time $\delta_1^v$ and rest for time $\delta_2^v$, respectively. Subsequently, the mosquito feeding rate is $f_R = \frac{1}{\delta_1^v + \delta_2^v}$. It is assumed that $\delta_2^v$ is unaffected by interventions such as ITNs or IRS, whilst the time searching for a bloodmeal increases to $\delta_1^v = \frac{\delta_{10}^v}{1-Z}$. Here, $\delta_{10}^v$ is the duration of foraging in the absence of indoor interventions such as ITNs or IRS.

Each mosquito survives the period of feeding and resting with probability $p_1$ and $p_2$. In the absence of interventions, the probability of a mosquito feeding is given by $p_{10} = \exp(-\mu_0 \delta_{10})$ and of resting is given by $p_2 = \exp(-\mu_0 \delta_2)$. Here, $\mu_0$ is the natural death rate of mosquitoes. In the presence of interventions, $p_2$ is unchanged but $p_1 = \frac{p_{10} w}{1 - Z p_{10}}$.

The probability of surviving a feeding cycle is $p_1^v p_2^v = \exp\left(\frac{-\mu^v}{f_R^v}\right)$. The mosquito death rate $\mu$ is then given by the following equation:

$$\mu = -f_R \log(p_1 p_2) \tag{28}$$

The force of infection from humans to vectors is given by the following equation, where $c_i$ is the infectivity of the infectious state that a person is in:

$$\Lambda^v = \sum_i c_i \lambda_i \tag{29}$$

The EIR experienced by each person due to the mosquito is $\lambda_i I_v$, where $I_v$ is the number of infectious vectors. The total experienced by people is the sum of this over all species. In this analysis, we only model 1 species: an *Anopheles gambiae*-like vector with a human blood index of 0.92 [3].

A phase II laboratory trial showed that ivermectin has a killing effect for approximately 23 days [14]. However, the blood-drug concentration wanes over time, thus reducing the efficacy of ivermectin depending on the time of the mosquito bloodmeal in relation to when the person was treated (S4 Fig). In the previously developed model with ivermectin MDA [10], there are 23 sets of Susceptible (S), pre-infectious (E) and infectious (I) mosquito compartments. Mosquitoes enter one of these sets of compartments depending on the time of the ivermectin bloodmeal in relation to when the person received the MDA. In each compartment, the hazard ratio on day $j$ since the MDA is used to inflate the mosquito mortality rate (30 and corresponds to that presented in the hazard ratio curve.

**Fig S4. Daily hazard ratios for mosquito death the 3x300$\mu$/kg dosage of ivermectin, from a phase II trial [14].** In this dosage, individuals are treated with ivermectin for 3 consecutive days, causing the blood-drug concentration to rise and subsequently, the hazard ratio to rise between day 1 and 3. The hazard ratios are consistently above 1 for 23 days. The horizontal dashed line corresponds to the hazard ratio of 1, where there is no increased probability of death mosquitoes ingest ivermectin compared to if they do not.

$$\mu_v^j = \mathrm{HR}^j \mu_v \tag{30}$$

Given the adaptations made to simulate slower MDA implementation campaigns, the hazard ratios and time-varying coverage ($Q_{lethal}(t)$) used to parameterise the model are shown in S1 Fig.

**Fig S5. Population-level hazard ratios and time-varying proportion of people with a toxic dose of ivermectin, used for simulating staggered and overnight distributions of ivermectin MDA, taking place monthly for three months**. Here, staggered distributions consist of three evenly interspaced distributions (akin to treating 3 groups of the total population) over a 10-day or 20-day period, whereas overnight distributions are completed in a single day. A: Hazard ratio curves for 3x300$\mu$g/kg dosage of ivermectin [14] were modified to account for staggered distributions. On each day, the average of hazard ratios in each group that were above 1 was taken to generate a new hazard ratio curve (shown by the black solid line) and used to parameterise a malaria transmission model which incorporates ivermectin MDA [10, 12]. The horizontal dashed line denotes a hazard ratio of 1, at which ivermectin has no additional killing effect on mosquitoes. B: At each time point, the proportion of covered groups with a hazard ratio greater than 1 ($K_{HR>1}(t)$. C: The population (%) with a lethal dose of ivermectin, estimated by multiplying the daily proportion of covered groups with the hazard ratio great than 1 (in panel B) by the covered population that are eligible for the MDA (using Equations 7 and 8 in the main text). The black solid line shows the proportion of the population with a lethal dose, if a target 70% of the population are treated (children under 5-years-old are not eligible for ivermectin, hence the black line never reaches 70%. The blue ribbon shows how the population with a lethal dose (%) varies depending on target coverage (lower bound: 50% target coverage; upper bound: 90% target coverage).

The differential equations governing this mechanistic model are given as follows:

Supporting Information Equation 31 is used to define the time when the endectocide is effective and when to move vectors to a new compartment, where t is time, $\tau$ is the time the administration starts and d is the number of days after the start of the endectocide administration and $L_{endec}$ is the duration of the killing effect of the endectocide, assumed here to be 23 days for an ivermectin-like endectocide:

$$\delta(t,\tau,d) = \begin{cases} 1 & \textit{if } \tau \leq t < \tau + d \\ 0 & \text{otherwise} \end{cases} \tag{31}$$

The rate at which mosquitoes feed on potentially treated host is given by $\alpha$, which is the product of the mosquito human blood index (here, assumed to be 0.92 for an *Anopheles-gambiae*-like mosquito) and the mosquito biting rate (it is assumed that mosquitoes take a bloodmeal once every 3 days).

Susceptible mosquitoes that have never bloodfed ($\mu_v$ is the mosquito mortality rate in the presence of indoor vector control interventions (ITNs or IRS), although here it is assumed that no indoor vector control interventions are in place, thus $\mu_v = \mu_{v0}$):

$$\frac{dS_v}{dt} = \beta - \mu_v S_v - \alpha S_v \tag{32}$$

where $\beta$ is given by the following ($m_{v0}$ is the vector-host ratio at endemic equilibrium and $\mu_{v0}$ is the natural mosquito mortality rate, here assumed to follow an exponential distribution with a mean of 0.1 deaths per day), and $\theta$ is a fourier function of changes in vector-host ratio given daily changes in rainfall within a year:

$$\beta = m_{v0}\mu_{v0}\theta \tag{33}$$

Susceptible mosquitoes that have bloodfed once:

$$\frac{dS^v_{F_1}}{dt} = \begin{cases} (\alpha - \Lambda^v)(1 - C_{endec})S^v - \alpha S^v_{F_1} - \mu^v_t S^v_{F_1} & \textit{if } \delta(t, T_{endec}, L_{endec}) = 1 \\ (\alpha - \Lambda^v)S^v - \alpha S^v_{F_1} - \mu^v_t S^v_{F_1} & \textit{if } \delta(t, T_{endec}, L_{endec}) = 0 \end{cases} \tag{34}$$

Susceptible mosquitoes that have bloodfed twice:

$$\frac{dS^v_{F_2}}{dt} = \begin{cases} (\alpha - \Lambda^v)(1 - C_{endec})S^v_{F_1} - (\Lambda^v(1 - C_{endec}) + \alpha C_{endec})\, S^v_{F_2} - \mu^v_t S^v_{F_2} & \textit{if } \delta(t, T_{endec} + j - 1, 1) = 1 \\ (\alpha - \Lambda^v)S^v_{F_1} - \Lambda^v S^v_{F_2} - \mu^v_t S^v_{F_2} & \textit{if } \delta(t, T_{endec}, L_{endec}) = 0 \end{cases} \tag{35}$$

Susceptible mosquitoes that have bloodfed once and acquired a bloodmeal containing endectocide on day $j$ after endectocide administration, where $j = 1,..L_{endec}$:

$$\frac{dS^{v,j}_{X,F_1}}{dt} = \begin{cases} C_{endec}(\alpha - \Lambda^v)S^v - \alpha S^{v,j}_{X,F_1} - \mu^{v,j} S^{v,j}_{X,F_1} & \textit{if } \delta(t, T_{endec} + j - 1, 1) = 1 \\ -\mu^{v,j} S^{v,j}_{X,F_1} - \alpha S^{v,j}_{X,F_1} & \textit{otherwise} \end{cases} \tag{36}$$

Susceptible mosquitoes that have bloodfed twice and acquired a bloodmeal containing endectocide on day

$j$ after endectocide administration, where $j$ = 1,...$L_{endec}$:

$$\frac{dS^{v,j}_{X,F_2}}{dt} = \begin{cases} C_{endec}(\alpha - \Lambda^v)(S^v_{F1} + S^v_{F2}) + (\alpha - \Lambda^v)S^{v,j}_{X,F_1} - \Lambda^v S^{v,j}_{X,F_1} - \mu^{v,j} S^{v,j}_{X,F_2} & \textit{if } \delta(t, T_{endec} + j - 1, 1) = 1 \\ (\alpha - \Lambda^v)S^{v,j}_{X,F_1} - \mu^{v,j} S^{v,j}_{X,F_2} - \Lambda^v S^{v,j}_{X,F_2} & \textit{otherwise} \end{cases} \quad (37)$$

Exposed (infected) mosquitoes that have bloodfed once and are on their $i$th day on the sporogenic cycle, where $i$ = 1 only:

$$\frac{dE^{v,i}_{F_1}}{dt} = \begin{cases} \Lambda^v(1 - C_{endec})S^v - \alpha E^{v,i}_{F_1} - \frac{1}{\tau^v} E^{v,i}_{F_1} - \mu^v_t E^{v,i}_{F_1} & \textit{if } \delta(t, T_{endec}, L_{endec}) = 1 \\ \Lambda^v S^v - \alpha E^{v,i}_{F_1} - \frac{1}{\tau^v} E^{v,i}_{F_1} - \mu^v_t E^{v,i}_{v,F_1} & \textit{if } \delta(t, T_{endec}, L_{endec}) = 0 \end{cases} \quad (38)$$

Exposed (infected) mosquitoes that have bloodfed once and are on their $i$th day on the sporogenic cycle, where i = 2,...10:

$$\frac{dE^{v,i}_{F_1}}{dt} = \frac{1}{\tau^v} E^{v,i-1}_{F_1} - \alpha E^{v,i}_{F_1} - \frac{1}{\tau^v} E^{v,i}_{F_1} - \mu^v_t E^{v,i}_{F_1} \quad (39)$$

Exposed (infected) mosquitoes that have bloodfed twice and are on their $i$th day on the sporogonic cycle, where $i$ = 1 only:

$$\frac{dE^{v,i}_{F_2}}{dt} = \begin{cases} \alpha(1 - C_{endec})E^{v,i}_{F_1} + \Lambda^v(1 - C_{endec})(S^v_{F_1} + S^v_{F_2}) - \alpha C_{endec} E^{v,i}_{F_2} - \frac{1}{\tau^v} E^{v,i}_{F2} - \mu^v_t E^{v,i}_{F_2} & \textit{if } \delta(t, T_{endec}, L_{endec}) = 1 \\ \alpha E^{v,i}_{F_1} + \Lambda^v(S^v_{F_1} + S^v_{F_2}) - \frac{1}{\tau^v} E^{v,i}_{F_2} - \mu^v_t E^{v,i}_{F_2} & \textit{if } \delta(t, T_{endec}, L_{endec}) = 0 \end{cases} \quad (40)$$

Exposed (infected) mosquitoes that have bloodfed twice and are on their $i$th day on the sporogonic cycle where i = 2,...10:

$$\frac{dE^{v,i}_{F_2}}{dt} = \begin{cases} \frac{1}{\tau^v} E^{v,i-1}_{F_2} + \alpha(1 - C_{endec})E^{v,i}_{F_1} - \alpha C_{endec} E^{v,i}_{F_2} - \frac{1}{\tau^v} E^{v,i}_{F_2} - \mu^v_t E^{v,i}_{F_2} & \textit{if } \delta(t, T_{endec}, L_{endec}) = 1 \\ \frac{1}{\tau^v} E^{v,i-1}_{F_2} + \alpha E^{v,i}_{F_1} - \frac{1}{\tau^v} E^{v,i}_{F_2} - \mu^v_t E^{v,i}_{F_2} & \textit{if } \delta(t, T_{endec}, L_{endec}) = 0 \end{cases} \quad (41)$$

Exposed (infected) mosquitoes that have bloodfed once and are on their ith day on the sporogonic cycle, where i = 1 only, and acquired a bloodmeal containing endectocide on day j after endectocide administration, where j = 1...$L_{endec}$:

$$\frac{dE^{v,i,j}_{X,F_1}}{dt} = \begin{cases} \Lambda^v C_{endec} S^v - \frac{1}{\tau^v} E^{v,i,j}_{X,F_1} - \alpha E^{v,i,j}_{X,F_1} - \mu^{v,j} E^{v,i,j}_{X,F_1} & \textit{if } \delta(t, T_{endec} + j - 1, 1) = 1 \\ -\frac{1}{\tau^v} E^{v,i,j}_{X,F_1} - \alpha E^{v,i,j}_{X,F_1} - \mu^{v,j} E^{v,i,j}_{X,F_1} & \textit{otherwise} \end{cases} \tag{42}$$

Exposed (infected) mosquitoes that have bloodfed once and are on the ith day of the sporogonic cycle, where i = 2...,10 only, and acquired a bloodmeal containing ivermectin on day j after ivermectin administration, where j = 1,...$L_{endec}$

$$\frac{dE^{v,i,j}_{X,F_1}}{dt} = \begin{cases} \frac{1}{\tau^v} E^{v,i-1,j}_{X,F_1} - \frac{1}{\tau^v} E^{v,i,j}_{X,F_1} - \alpha E^{v,i,j}_{X,F_1} - \mu^{v,j} E^{v,i,j}_{X,F_1} & \textit{if } \delta(t, T_{endec} + j - 1, 1) = 1 \\ \frac{1}{\tau^v} E^{v,i-1,j}_{X,F_1} - \frac{1}{\tau^v} E^{v,i,j}_{X,F_1} - \alpha E^{v,i,j}_{X,F_1} - \mu^{v,j} E^{v,i,j}_{X,F_1} & \textit{otherwise} \end{cases} \tag{43}$$

Exposed (infected) mosquitoes that have bloodfed twice and are on their ith day on the sporogonic cycle, where i = 1 only, and acquired a bloodmeal containing endectocide on day j after endectocide administration, where j = 1,..Lendec:

$$\frac{dE^{v,i,j}_{X,F_2}}{dt} = \begin{cases} \Lambda^v C_{endec}(S^v_{F1} + S^v_{F2}) + \Lambda^v (S^{v,j}_{X,F1} + S^{v,j}_{X,F2}) + & \\ \alpha C_{endec}(E^{v,i}_{F1} + E^{v,i}_{F2}) + \alpha E^{v,i,j}_{X,F1} - \frac{1}{\tau^v} E^{v,i,j}_{X,F2} - \mu^{v,j} E^{v,i,j}_{X,F2} & \textit{if } \delta(t, T_{endec} + j - 1, 1) = 1 \\ \alpha E^{v,i,j}_{X,F1} + \Lambda^v (S^{v,j}_{X,F1} + S^{v,j}_{X,F2}) - \frac{1}{\tau_v} E^{v,i,j}_{X,F2} - \mu^{v,j} E^{v,i,j}_{X,F2} & \textit{otherwise} \end{cases} \tag{44}$$

Exposed (infected) mosquitoes that have bloodfed twice and are on their ith day on the sporogonic cycle, where i = 2,..10 only, and acquired a bloodmeal containing endectocide on day j after endectocide administration, where j = 1,...$L_{endec}$:

$$\frac{dE^{v,i,j}_{X,F_2}}{dt} = \begin{cases} \frac{1}{\tau^v} E^{v,i-1,j}_{X,F2} + \alpha C_{endec}(E^{v,i}_{F1} + E^{v,i}_{F2}) + \alpha E^{i,j}_{v,X,F1} - \frac{1}{\tau^v} E^{v,i,j}_{X,F2} - \mu^{v,j} E^{v,i,j}_{X,F2} & \textit{if } \delta(t, T_{endec} + j - 1, 1) = 1 \\ \alpha E^{v,i,j}_{X,F1} + \frac{1}{\tau^v} E^{v,i-1,j}_{X,F2} - \frac{1}{\tau^v} E^{v,i,j}_{X,F2} - \mu^{v,j} E^{v,i,j}_{X,F2} & \textit{otherwise} \end{cases} \tag{45}$$

Infectious mosquitoes that have bloodfed once:

$$\frac{dI^v_{F1}}{dt} = \frac{1}{\tau^v} E^{v,10}_{F1} - \alpha I^v_{F1} - \mu^v_t I^v_{F1} \tag{46}$$

Infectious mosquitoes that have bloodfed twice:

$$\frac{dI^v_{F2}}{dt} = \begin{cases} \frac{1}{\tau^v}E^{v,10}_{F2} + \alpha(1-C_{endec})I^v_{F1} - \alpha C_{endec}I^v_{F2} - \mu^v_t I^v_{F2} & \textit{if } \delta(t, T_{endec}, L_{endec}) = 1 \\ \frac{1}{\tau^v}E^{v,10}_{F2} + \alpha I^v_{F1} - \mu^v_t I^v_{F2} & \textit{otherwise} \end{cases} \tag{47}$$

Infectious mosquitoes that have bloodfed once and acquired a bloodmeal containing endectocide on day j after endectocide administration, where j = 1,...$L_{endec}$:

$$\frac{dI^v_{X,F1}}{dt} = \frac{1}{\tau^v}E^{v,10,j}_{X,F1} - \alpha I^{v,j}_{X,F1} - \mu^{v,j} I^{v,j}_{X,F1} \tag{48}$$

Infectious mosquitoes that have bloodfed twice and acquired a bloodmeal containing endectocide on day j after endectocide administration, where j = 1,...$L_{endec}$:

$$\frac{dI^v_{X,F2}}{dt} = \begin{cases} \alpha C_{endec}(I^v_{F1} + I^v_{F2}) + \frac{1}{\tau^v}E^{v,10,j}_{X,F2} + \alpha I^{v,j}_{X,F1} - \mu^j_v I^{v,j}_{X,F2} & \textit{if } \delta(t, T_{endec} + j - 1, 1) = 1 \\ \frac{1}{\tau^v}E^{v,10,j}_{X,F2} + \alpha I^{v,j}_{X,F1} - \mu^{v,j} I^{v,j}_{X,F2} & \textit{otherwise} \end{cases} \tag{49}$$

In this theoretical modelling study, we assume malaria transmission is by an *Anopheles gambiae*-like vector (with a high human blood index of 0.92) and that no other vector control interventions are present in the population. We simulate transmission in perennial and seasonal high transmission setting (initial annual entomological inoculation rate, EIR, of 100 bites per person per year). S2 Fig shows the model-predicted dynamics in the daily EIR and clinical incidence in children under 5-years old, per 1000 persons, in the seasonal scenarios where the MDA is either on-time in relation to seasonality in mosquito abundance or 2 months later than planned, of which the latter could occur due to logistical constraints. S7- S9 Figs show these results in these settings with low endemicity (initial annual EIR of 2).

**Fig S6. Transmission dynamics (daily EIR per person and clinical incidence of malaria in children under 5-years-old in a high endemicity and highly seasonal setting, where MDA deployment of an ivermectin-like endectocide is either on-time, in relation to seasonality, or late, of which the latter can occur due to logistical constraints.** The late MDA campaigns are simulated to start 60 days later than the well-time MDA, coinciding with the time of the peak in clinical incidence. Panels A and C show the dynamics in the daily EIR (average number of infectious bites per person per day) in relation to the start of the on-time MDA. Panels B and D show the dynamics in clinical incidence of malaria in children under 5-years-old (per 1000 persons), in relation to the start of the on-time MDA. In panels A-D, the light green line shows a scenario where no interventions are introduced, whilst other colours show varying times for MDA deployment, be it the synchronised (blue), 10-day MDA (dark green) or 20-day MDA (orange). MDAs take place once a month for 3 months (denoted by the 3 black arrows). Panels A and B show model-predicted dynamics in the scenario where the MDA starts on-time in relation to seasonality in mosquito abundance, whereas panel C and D show dynamics where the MDa starts 60 days later than planned. The black box in panels B and D shows the 6-month time period, following the start of the MDA, over which efficacy is measured (as the percentage of cases averted by the endectocide), shown in the far left bars of Figure reffig3E. The blue shaded area shows the 1-year period, since the start of the MDA, over which which efficacy is measured (as the percentage of cases averted by the endectocide), shown in the far right bars of Figure reffig3E.

**Fig S7. Transmission dynamics and efficacy of synchronised or slow MDA deployments of ivermectin-like endectocide in a low endemicity and perennial setting.** Panel A-D show, respectively, mosquito density (vector-host ratio), daily entomological inoculation rate (EIR), malaria all-age slide prevalence (%) and clinical incidence of malaria in children under 5-years-old. Summary efficacy estimates are shown in Panel E. In Panels A-D, the light green line shows a scenario where no interventions are introduced, whilst the other colours show varying times for MDA deployment, be it synchronised, i.e. all individuals due to receive the MDA do so on the same day, (blue), 10 days (dark green) or 20 days (orange). MDAs take place once a month for 3 months (denoted by the 3 black arrows). The dark blue arrow in panel C shows when prevalence is measured (1 month after the last MDA, efficacy shown in middle set of bars of panel E). In panel D the black rectangular and the blue shaded area show, respectively, the 6-month and 12-month periods over which the cumulative number of clinical cases averted was calculated (the % difference from the no intervention arm shown as the other efficacy bars in panel E) . Panel E: The efficacy of synchronised and slow deployments, measured by calculating the relative reduction (%) in clinical incidence in children under 5-years-old compared to a baseline scenario of no interventions. The height of the bars corresponds to the efficacy derived from a simulated endectocide coverage of 70% amongst eligibles, with the vertical error bars correspond to a 50% (lower bound) or 90% (upper bound) simulated endectocide coverage.

**Fig S8. Transmission dynamics and efficacy of synchronised or slow MDA deployments of ivermectin-like endectocide in a low endemicity and highly seasonal setting, where the timing of the MDA is deemed on-time in relation to seasonality in mosquito abundance.** Panel A-D show, respectively, mosquito density (vector-host ratio), daily entomological inoculation rate (EIR), malaria all-age slide prevalence (%) and clinical incidence of malaria in children under 5-years-old. Summary efficacy estimates are shown in Panel E. In Panels A-D, the light green line shows a scenario where no interventions are introduced, whilst the other colours show varying times for MDA deployment, be it synchronised, i.e. all individuals due to receive the MDA do so on the same day, (blue), 10 days (dark green) or 20 days (orange). MDAs take place once a month for 3 months (denoted by the 3 black arrows). The dark blue arrow in panel C shows when prevalence is measured (1 month after the last MDA, efficacy shown in middle set of bars of panel E). In panel D the black rectangular and the blue shaded area show, respectively, the 6-month and 12-month periods over which the cumulative number of clinical cases averted was calculated (the % difference from the no intervention arm shown as the other efficacy bars in panel E) . Panel E: The efficacy of synchronised and slow deployments, measured by calculating the relative reduction (%) in clinical incidence in children under 5-years-old compared to a baseline scenario of no interventions. The height of the bars corresponds to the efficacy derived from a simulated endectocide coverage of 70% amongst eligibles, with the vertical error bars correspond to a 50% (lower bound) or 90% (upper bound) simulated endectocide coverage.

**Fig S9. Transmission dynamics and efficacy of synchronised or slow MDA deployments of ivermectin-like endectocide in a low endemicity and highly seasonal setting, where the timing of the MDA is late in relation to seasonality in mosquito abundance.** The late MDA deployment starts 60 days later than planned, coinciding with the peak in clinical incidence. Panel A-D show, respectively, mosquito density (vector-host ratio), daily entomological inoculation rate (EIR), malaria all-age slide prevalence (%) and clinical incidence of malaria in children under 5-years-old. Summary efficacy estimates are shown in Panel E. In Panels A-D, the light green line shows a scenario where no interventions are introduced, whilst the other colours show varying times for MDA deployment, be it synchronised, i.e. all individuals due to receive the MDA do so on the same day, (blue), 10 days (dark green) or 20 days (orange). MDAs take place once a month for 3 months (denoted by the 3 black arrows). The dark blue arrow in panel C shows when prevalence is measured (1 month after the last MDA, efficacy shown in middle set of bars of panel E). In panel D the black rectangular and the blue shaded area show, respectively, the 6-month and 12-month periods over which the cumulative number of clinical cases averted was calculated (the % difference from the no intervention arm shown as the other efficacy bars in panel E) . Panel E: The efficacy of synchronised and slow deployments, measured by calculating the relative reduction (%) in clinical incidence in children under 5-years-old compared to a baseline scenario of no interventions. The height of the bars corresponds to the efficacy derived from a simulated endectocide coverage of 70% amongst eligibles, with the vertical error bars correspond to a 50% (lower bound) or 90% (upper bound) simulated endectocide coverage.
